# Quantifying optimal AWaRe antibiotic use patterns in hospitals with a new benchmarking tool applied to cross-sectional point prevalence survey data from 10 countries

**DOI:** 10.64898/2026.10.12.26363793

**Authors:** Myo Maung Maung Swe, Ines Pauwels, Silvia Figueiredo Costa, Ana Paula Matos Porto, Mari Rose De Los Reyes, Jemelyn Garcia, Jafar Soltani, David C Banda, Svetlana Rachina, Yuliya Belkova, Herberth Maldonado, Milena Lopicic, Maja Raicevic, Erika Vlieghe, Ann Versporten, Michael Sharland, Ben S Cooper, Cherry Lim, on behalf of the ADILA consortium and the Global-PPS network

## Abstract

**Background:** Both over- and under-antibiotic use pose major global health challenges, yet there is no consensus on how to quantify deviations from optimal prescribing or set evidence-based targets. We developed a benchmarking tool for inpatient antibiotic use and evaluated its application in 10 countries.

**Methods:** We developed decision-tree models to estimate expected empiric antibiotic use for 11 indications, based on the WHO AWaRe book for two age groups: 28 days to ≤12 years and >12 years. We parameterised the models using country-specific antimicrobial resistance estimates in the 10 countries and cross-sectional Global Point Prevalence Survey (Global-PPS) case-mix data from 2021 for 10 countries and from 2015–2022 in a subset of five countries. We then compared estimated expected antibiotic use with observed empiric use (from Global-PPS) in defined daily doses for adults and days of therapy for children, across indications and AWaRe categories.

**Findings:** In 2021, among patients aged >12 years, in all countries but one, observed empiric Access antibiotic use, as a proportion of total antibiotic use, was lower than expected and empiric Watch antibiotic use was higher than expected. Similar patterns were observed in all countries but one in patients aged 28 days to ≤12 years. The magnitude of discrepancies between expected and observed Access and Watch antibiotic use varied across indications and countries. The median observed-to-expected ratios of usage across countries ranged from 0.3–0.8 for Access antibiotics, and 0.7–5.3 for Watch antibiotics in patients aged >12 years in 2021. Temporal trends from repeated surveys from 2015 to 2022 showed little change in observed-to-expected ratios for Access and Watch antibiotics.

**Interpretation:** This benchmarking tool uses Global-PPS data to quantify differences between observed antibiotic use and WHO AWaRe recommendations. Our tool can be adaptable to local guidelines, provides policy-relevant estimates of expected antibiotic use to support monitoring, procurement planning, and targeted stewardship intervention.

**Funding:** This study is part of the ADILA project supported by the Wellcome Trust (222051/Z/20/Z). The Global-PPS is supported by bioMérieux. bioMérieux is the sole private partner of the Global-PPS and had no role or responsibility in the study design, data collection, data analysis, data interpretation or writing the report.

**Research in context:** *Evidence before this study:* Monitoring antimicrobial use and benchmarking observed against expected antimicrobial usage given local disease burden and prescribing guidelines has the potential to help optimise antimicrobial use. We searched PubMed on 3 August 2026 for articles published since 1 January 2020, using the search terms: (((Benchmark) AND (Expected)) AND (Antimicrobial)) AND (Hospital). Among 37 studies retrieved, we found eight that benchmarked observed antimicrobial use against an estimate of expected use in hospital settings for interhospital comparison, all of which were conducted in high-income countries (the United States, United Kingdom, Finland and Japan). A range of statistical approaches including indirect standardisation, risk-adjusted regression, and generalised linear mixed models were used to estimate expected antibiotic use. Notably, none of these studies estimated the expected use based on antibiotic treatment guidelines, drug availability and risk of antimicrobial resistance (AMR) in the study population.

*Added value of this study:* We developed a novel tool for estimating expected empirical antibiotic use based on the World Health Organization (WHO) guidelines while accounting for local disease burden, risk of antibiotic resistance, drug availability and patient severity. We developed decision tree models informed by the recommended empirical antibiotic treatment for ten common infection syndromes and surgical prophylaxis for hospital settings in the WHO’s AWaRe antibiotic book. The models were informed by data from the Global Point Prevalence Survey (Global-PPS) to estimate expected antibiotic use by AWaRe category in both relative (as a proportion of total use) and absolute measures. These estimates were used to benchmark the observed antibiotic use in the same cohort. We then calculated observed-to-expected ratios to quantify the deviation of the observed antibiotic use from the expected use, given reported local disease burden, and demonstrated the utility of these ratios for monitoring changes in antibiotic usage patterns over time. The findings indicated that Watch antibiotic use was substantially higher than would be expected if the surveyed hospitals had followed the WHO empirical antibiotic prescription guidelines, and there were improvements in Watch plus antibiotic usage for patients with age >12 years over the study period.

*Implications of all the available evidence:* This new analytical tool could enable hospitals to assess deviations in empiric antibiotic use relative to the expected use in both absolute terms (e.g. measures in defined daily doses and days of therapy) and proportional measures (out of the total antibiotic use) by AWaRe category. This tool can also be readily adaptable to incorporate local treatment guidelines. By identifying specific AWaRe categories and clinical indications where observed use deviates most from expectations, the framework provides a new tool to support the prioritisation of targeted antimicrobial stewardship interventions. Moreover, this tool could be used to monitor antibiotic use patterns in hospitals over time and identify possible cost savings.

## Introduction

Antibiotics are critical to modern medicine but there is ample evidence that their use (and over-use) is driving the emergence and spread of resistance, threatening their effectiveness.^1,2^ In 2017, the World Health Organization (WHO) established the AWaRe system as a tool to support antimicrobial stewardship.^3^ This system classifies antibiotics into three groups Access, Watch and Reserve, based on their potential impacts on emergence of antimicrobial resistance. An accompanying document, the WHO AWaRe antibiotic book, provides empiric treatment guidance for 34 common infections in primary care and hospital settings both for children and adults.^4^ In 2024, the United Nations General Assembly (UNGA) high-level meeting endorsed the AWaRe classification system as a standard method to monitor antibiotic consumption and agreed on a global target of Access antibiotics to be at least 70% of overall usage in humans by 2030.^5^ Establishing national targets for antibiotic consumption based on the local burden of infectious diseases and the associated risk of antimicrobial resistance (AMR) is necessary for producing context-specific antibiotic use recommendations.^6^

In recent years, growing evidence has documented increased use of broad-spectrum antibiotics including vancomycin, carbapenems, third-generation cephalosporins, and last-resort antibiotics in hospital settings.^7,8^ The empirical use of these antibiotics should be guided by the local burden and type of infection, severity of disease, risk of AMR, and treatment guidelines. However, an appropriate benchmarking framework to assess whether antibiotic use aligns with these factors is currently lacking. Existing approaches largely focus on benchmarking performance across facilities using routinely collected administrative or medical record data, primarily to identify outlier hospitals and support antimicrobial stewardship programmes, rather than to define context-specific, appropriate levels of antibiotic use.^9–16^

With the increasing availability of Point Prevalence Survey (PPS) study data in low- and middle-income countries, there is an opportunity to link hospital antibiotic usage data to local disease burden data, enabling a comparison between observed antibiotic use and that which would be expected had prescribing followed WHO AWaRe guidelines. Here we introduce such a benchmarking tool and apply it to PPS data from ten countries to determine the extent to which observed levels of antibiotic use by AWaRe classification differ from those which would be expected if prescribing followed the WHO guidelines.

## Methods

### Decision tree model

We developed two decision tree models: one for patients aged over 12 years (hereafter referred to as patients aged >12 years), and one for patients aged from 28 days to 12 years (hereafter referred to as patients aged ≤12 years), according to the empirical treatment guidelines in the WHO AWaRe Antibiotic Book.^4^ The WHO AWaRe Antibiotic Book was used as a case example to demonstrate how our approach could directly account for an existing prescription guideline. Neonates were excluded from the analysis because of lack of clinical guidance. We modelled eleven indications, including ten infection syndromes and surgical prophylaxis. The ten infection syndromes were community-acquired pneumonia, hospital-acquired pneumonia (non-ventilator associated), bacterial meningitis, intra-abdominal infection (acute cholecystitis and cholangitis, pyogenic liver abscess, acute appendicitis, acute diverticulitis), acute pyelonephritis, skin and soft-tissue infection (pyomyositis and necrotizing fasciitis), bone and joint infection (acute bacterial osteomyelitis and septic arthritis), *Clostridioides difficile* infection, febrile neutropenia, and sepsis (and septic shock).

For indications where the guidelines suggested two different treatment options based on severity, the tree included two branches to account for severe and non-severe cases. When the guideline further differentiated treatment options within each severity level into first- and second-choice, we added a daughter node representing the likelihood of receiving either option. For indications with multiple antibiotic options under each choice recommended by the AWaRe guidelines, we included additional branches to allow for those options. For infections where the guideline accounted for the risk of AMR infection, for example treatment recommendations accounted for local burden of extended spectrum beta-lactamase producing bacteria (ESBL) or methicillin-resistant *Staphylococcus aureus* (MRSA), additional branches were added to factor in the local burden of AMR infections. The complete decision trees can be found in Supplementary Figure 1 - 11. All analyses were performed using *R* (version 4.2.0; The R Foundation for Statistical Computing, Vienna, Austria). Trees were visualised using *Silver Decisions 1.2.0* (https://silverdecisions.pl/).^17^

### Populating the model

#### Data sources

We used Global-PPS data to populate the model and to calculate observed antibiotic usage. This is hospital antibiotic prescribing data from a 1-day point prevalence survey on inpatient hospital wards, collected according to the Global-PPS protocol.^18^ Participation in the Global-PPS project by hospitals was voluntary, and local ethical approval was obtained where necessary. All data were pseudonymised at the patient level. The Global-PPS data collection methods have been described in detail elsewhere.^19^ Two datasets from the Global-PPS were analysed to generate input parameters for the decision tree models. The first dataset contained data from ten countries (Brazil, Guatemala, India, Iran, Mexico, Montenegro, Nigeria, the Philippines, the Russian Federation and Zambia) in 2021, and the second dataset contained data from five countries (Brazil, Iran, the Philippines, the Russian Federation and Nigeria) from 2015 to 2022.

To calculate observed antibiotic use, for the cohort of patients aged >12 years, the prescribed dose on the day of the PPS in DDDs was calculated based on dosing and frequency and the WHO Anatomical Therapeutic Chemical (ATC) classification system for each antibiotic prescription.^20^ For the cohort of patients aged ≤12 years, days of therapy (DOTs) was calculated. Only prescriptions for empirical antibiotics were included in this study. Details on calculation of DDDs and DOTs are reported in the Supplementary materials.

Global-PPS data used to generate input parameters for the decision tree models included the number of patients on antibiotics at the time of the PPS with different clinical infections, and types of surgery where surgical prophylaxis was indicated. A prescription was classified as targeted if assessable microbiological results were available on the day of the survey, and as empiric otherwise. Only the empiric antibiotic treatments were analysed and used to populate the models in this study.

Country- and year-specific prevalence estimates for two resistant bacteria considered in the WHO AWaRe antibiotic book when recommending empirical treatment — ESBL (Extended Spectrum Beta Lactamase) producing *Enterobacterales* and MRSA (methicillin resistant *Staphylococcus aureus*) were obtained from the WHO GLASS AMR dashboard ^21^. When year-specific AMR data were unavailable, data from the most recent previous year for the same country was used. Where country-specific AMR data were unavailable on the WHO GLASS dashboard, we used AMR estimates from the Global Research on Antimicrobial Resistance study^22^. Further details are provided in Supplementary table 1.

#### Probability distributions for key parameters in the decision trees

Uncertainty in probability parameters was represented using beta distributions for binary outcomes and Dirichlet distributions for multinomial outcomes to constrain the parameter value between zero and one. The beta distribution parameters included the proportion of patients with a given clinical condition receiving first choice treatment, the proportion presenting with severe disease, and the probability of AMR infection. The parameters of the Dirichlet distribution included the probabilities of underlying causes of sepsis (unknown origin, meningitis, lower respiratory tract infection, enteric fever, intra-abdominal infection, skin and soft tissue infection and urinary tract infection) in patients >12 years, and the probabilities of four different types of surgical prophylaxis in both age groups. Detailed information on the model parameters with corresponding probability distributions and justifications for the selected values are shown in the Supplementary Table 1.

#### Assumptions and scenarios modelled

We considered two scenarios, varying parameters related to the proportion of patients with a given clinical condition presenting with severe disease. In Scenario 1, the proportion of ICU admissions among surveyed patients was used as the input parameter representing severe disease in the study population. In Scenario 2, the proportion of cases receiving at least one parenteral antibiotic among surveyed patients was used as the input parameter for severe disease in the study population. In both scenarios, a hospital was defined as a high-risk setting for ESBL infection if the reported country- and year-specific proportion of ESBL among *Enterobacterales* infections exceeded 20%. Broad-spectrum antibiotics (e.g., meropenem) are expected to be used empirically in high-risk or severe cases of intra-abdominal infection, febrile neutropenia, and sepsis in accordance with the WHO AwaRe antibiotic book. We further categorised Watch antibiotics into two groups: Watch plus, which consisted of carbapenems and vancomycin; and Watch, which consisted of all other antibiotic classes within the Watch group.

#### Analysis

We performed a probabilistic analysis by propagating uncertainty through the decision tree model to reflect uncertainty associated with the parameter values. This was achieved by iterating the model 1,000 times, with each iteration sampling parameter values from their respective probability distributions. Each iteration produced a sample of both the total and syndrome-specific expected empirical antibiotic use classified by AWaRe category in DDD and DOT for patients aged >12 years and ≤12 years, respectively. These estimates were expressed as proportions of expected antibiotic use relative to total expected antibiotic use. The 95% uncertainty intervals (UI) for the expected antibiotic use relative to total antibiotic use were derived from the 2.5^th^ and 97.5^th^ percentiles of distributions. The observed antibiotic use, calculated in DDD and DOT from the Global-PPS data, was expressed as proportions of observed antibiotic use relative to total observed antibiotic use recorded, by AWaRe category and indication. To quantify the deviation of observed antibiotic use from the expected use, we calculated the ratios of observed-to-expected use from the absolute DDD and DOT values for patients aged >12 years and ≤12 years, respectively. Ratios derived from the 2021 data were visualised using heatmaps. To illustrate the utility of the observed-to-expected antibiotic use ratio for monitoring temporal changes, we fitted a Bayesian multilevel model to the ratios derived from the 2015 to 2022 data. These ratios were modelled using a truncated normal distribution, with a nonlinear effect of year that varied by AWaRe category, a random intercept for country, a random intercept for AWaRe category within each country, and a temporal autocorrelation term to account for the time-series structure of the data.

## Results

We analysed Global-PPS data from 7119 patients aged >12 years, representing 10,475 DDDs of antibiotic use and 1244 patients aged ≤12 years, representing 1830 DOTs, from participating hospitals in ten countries in 2021. We also analysed data from repeated cross-sectional surveys conducted in five countries between 2015 and 2022, comprising 25,475 patients aged >12 years (36,196 DDD) and 5162 patients aged ≤12 years (7278 DOT).

### Observed and expected antibiotic use as proportions of total observed use in 2021

Among patients aged >12 years, the proportion of expected antibiotic use out of the total use varied across countries by AWaRe category under both scenarios, with Access antibiotics constituting the largest proportion, followed by Watch and Watch plus antibiotics (Figure 1). In Scenario 1, using the proportion of ICU admissions among surveyed patients to represent severe cases in the study population, Access antibiotics were expected to account for at least 60% of the total use across all countries and the expected use of Watch and Watch plus antibiotics was below 30% and 20%, respectively, except in India. The observed proportion of Access antibiotic use out of the total was lower than expected, whereas Watch was higher than expected in all countries except in India. All countries had a higher observed proportion of Watch plus antibiotic use than expected. However, the small number of surveyed patients from the PPS data from study sites in Brazil, India, Mexico and Zambia resulted in wide uncertainty intervals around the expected antibiotic use. The observed use of Access antibiotics accounted for 20% to 30% of the total use in Brazil, Mexico, the Philippines and the Russian Federation, despite their expected use being between 60% and 85%. In contrast, reported Watch antibiotic use accounted for 60% to 75 % of the total use in Brazil, the Philippines, the Russian Federation and Zambia, despite their expected use lying between 10% to 30%. No use of Watch plus antibiotics was reported from the Global-PPS data in Zambia. The reported proportion of Watch plus antibiotic use in Nigeria aligned with the expected estimate.

**Figure 1.**
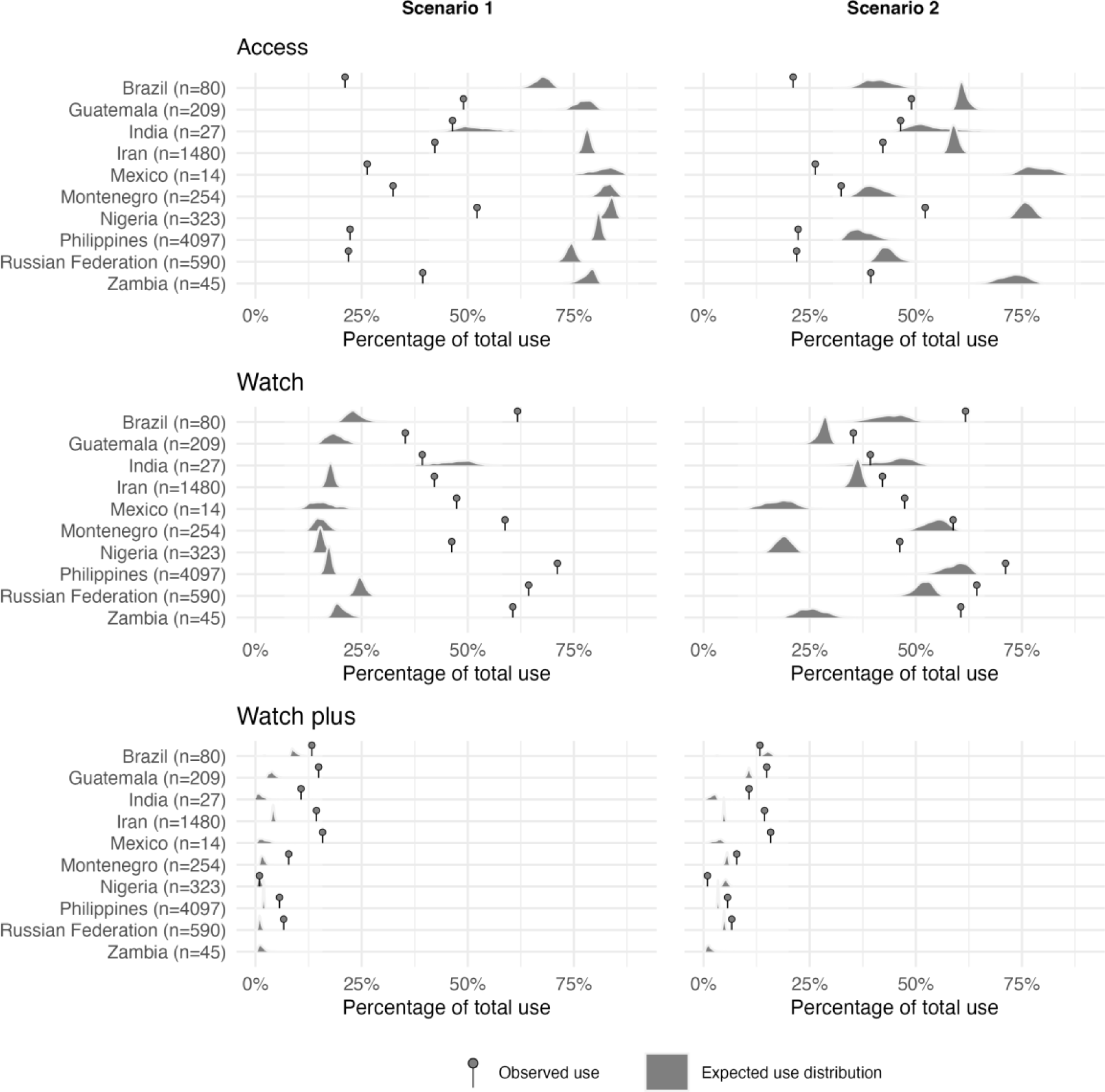
The proportions of observed and expected antibiotic use relative to total antibiotic use across the three AWaRe categories in patients >12 years. Lollipop markers indicate the observed proportion of antibiotic use, defined as the observed use of Access, Watch, and Watch plus antibiotics relative to total observed antibiotic use. Density plots represent the expected distributions of antibiotic use across AWaRe categories related to total expected antibiotic use. Both numerators and denominators were based on antibiotic use measured in defined daily doses (DDD) in 2021. The total number of patients on empirical treatment used to estimate expected antibiotic use is shown in brackets (n).

In Scenario 2, in which the proportion of cases receiving at least one parenteral antibiotic among surveyed patients was used to represent severe disease, the estimated proportion of expected Access antibiotic use was lower, and those of Watch and Watch plus antibiotics were higher than in Scenario 1. This suggests that in settings with a high proportion of severe disease cases, the gap between observed and expected use could be narrower than in settings where this proportion was low. Nevertheless, across all countries, the proportion of observed Access antibiotic use was consistently lower than the expected use and the observed Watch antibiotic use was consistently above the expected use in all countries apart from India. A similar pattern was observed in patients aged ≤ 12 years in both scenarios in 2021 (Supplementary Figure 15).

### Differences between expected and observed antibiotic use by indication

Surgical prophylaxis was the most prevalent indication of antibiotic use in Guatemala, India, Iran, Montenegro and Nigeria among patients aged >12 years (Supplementary Figure 12). In patients aged ≤12 years, the most common indication of antibiotic use was community-acquired pneumonia in the Philippines, Guatemala, Nigeria and Iran (Supplementary Figure 12).

Under Scenario 1, the indication with the largest underuse of Access antibiotics compared to expected use (relative to total use for each indication) in patients aged >12 years varied by country: bone and joint infections in Brazil, Iran, Nigeria, and the Russian Federation, community-acquired pneumonia in Mexico and the Philippines, hospital-acquired pneumonia in Guatemala, surgical prophylaxis in Zambia, and skin and soft tissue infections in India and Montenegro (Figure 2 and Supplementary Figure 16-17). Interestingly, among sepsis patients aged >12 years, the observed proportion of Watch antibiotic use was substantially lower than expected in participating hospitals in Brazil, Guatemala, the Russian Federation, and Iran. The expected proportions were 56.7% (95% UI: 53.6–62.6%), 56.3% (95% UI: 53.1–62.3%), 57.4% (95% UI: 54.5–61.1%), and 58.2% (95% UI: 57.0–59.5%), respectively, compared with observed proportions of 21.4%, 22.6%, 29.1%, and 35.1%. Conversely, for Watch plus antibiotics, the proportion of observed use was higher than expected. A similar pattern was also observed under Scenario 2, though in this case the discrepancies between expected and observed proportions of antibiotics in different AWaRe categories were smaller (Supplementary Figure 18-19).

**Figure 2.**
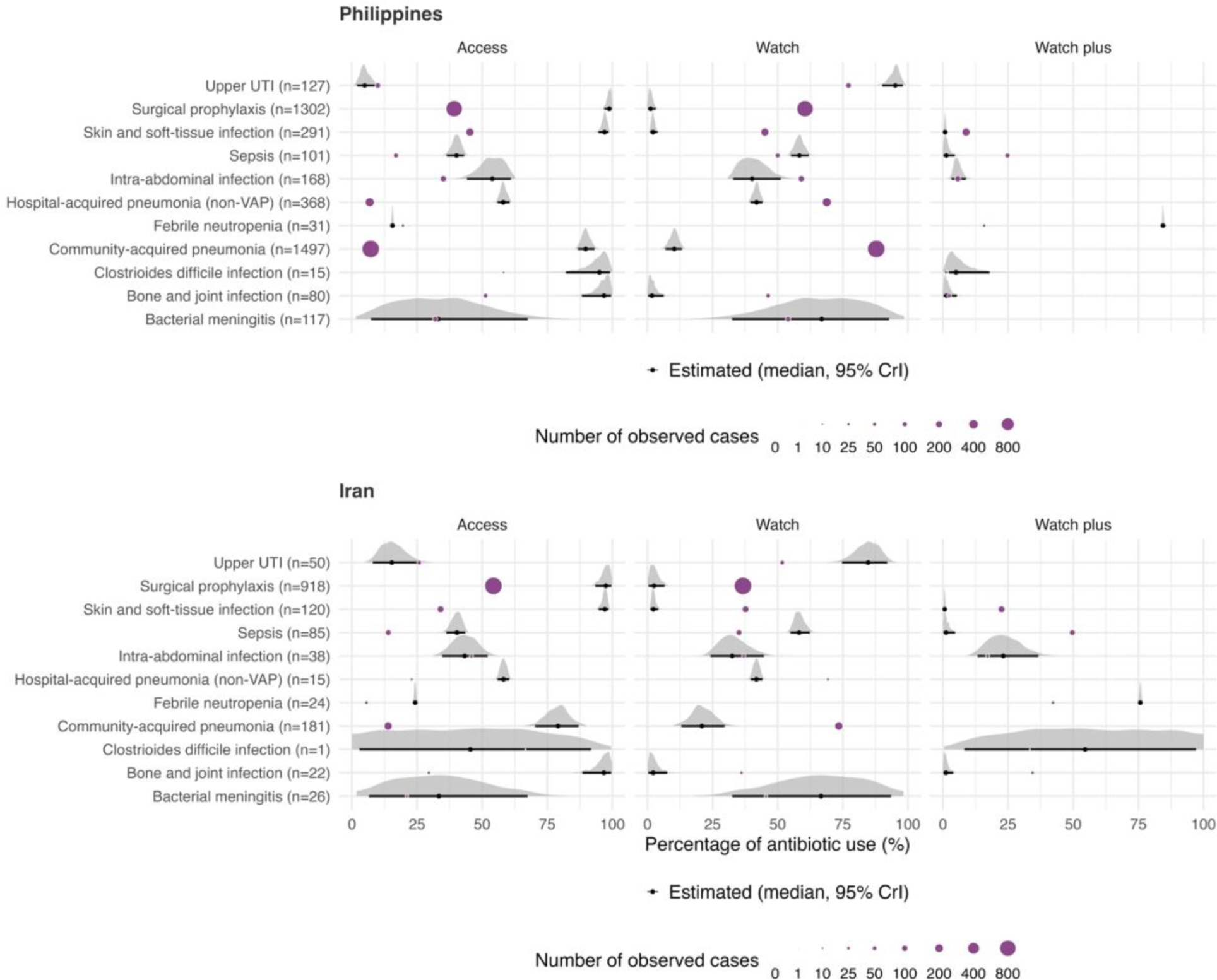
Proportion of observed and expected empiric antibiotic use out of the total use within each indication under Scenario 1 in patients aged >12 years in 2021 in the Philippines and Iran. Purple coloured circular dots indicate the proportion of observed use within each indication. The ridge line plot represents the distribution of the proportion of the expected use. The total number of patients on empirical treatment used to estimate expected antibiotic use is shown in brackets (n). UTI: urinary tract infection; VAP: ventilator-associated pneumonia.

Our benchmarking tool can be used to identify indications where prescribing interventions might be expected to have the largest impacts in reducing the overuse of Watch antibiotics. For instance, in the Philippines, the benchmarking tool suggests surgical prophylaxis and community-acquired pneumonia as indications where prescribing interventions should be targeted (Figure 2). If the participating hospitals in the Philippines had adhered to the WHO AWaRe guidelines, there would have been a modest reduction in total antibiotic use from 5912 DDDs to 5357 (95% UI: 5234 – 5603) but a major reduction in Watch antibiotic use from 4205 to 925 (95% UI: 883 – 973), with Watch plus use decreasing from 334 to 102 (95% UI: 92.2 – 114.7). In contrast, use of Access antibiotics would be expected to increase from 1316 to 4329 (95% UI: 4187.1 – 4587.1). In Iran, the benchmarking tool suggests that targeting antibiotic use for surgical prophylaxis would be one of the most impactful ways of reducing inappropriate Watch antibiotic use (Figure 2). If participating hospitals in Iran had adhered to the WHO AWaRe guidelines for such prophylactic use, the total antibiotic use would have been reduced from the observed 2327 DDDs to 1693 DDDs (95% UI: 1676.2 – 1718.0) with Watch antibiotic use decreasing from 980 to 299 (95% UI: 273 – 327), Watch plus antibiotic use decreasing from 334 to 71 (95% UI: 65 – 80) and Access antibiotic use increasing from 983 to 1324 (95% UI: 1303 – 1342). I

### Observed-to-expected use ratios based on absolute measure of antibiotic usage

The observed-to-expected use ratios quantify the magnitude of the discrepancy in absolute measures between observed and expected antibiotic use. The discrepancy between observed and expected antibiotic use was more pronounced for Watch Plus antibiotics compared with the discrepancies observed for Watch antibiotics in 2021 across most countries (Figure 3). In Scenario 1, observed absolute use of Watch antibiotics was between 2 to 5 times higher than expected in patients aged >12 years, except for Watch antibiotic use in India. For Watch plus antibiotic use, the observed antibiotic use was 3 to 8 times higher than expected, except in Brazil and Nigeria, where observed use aligned with estimated expected use accounting for the reported disease burden. In Scenario 2, the estimated observed-to-expected ratio for Watch plus antibiotic was 0.2 (95% UI, 0.2–0.3) among patients aged >12 years in Nigeria. This lower-than-expected use of empiric Watch plus antibiotics was observed for bone and joint infections, intra-abdominal infections, sepsis and skin and soft-tissue infections under assumptions of a high proportion of MRSA and ESBL pathogens and a high proportion of severe cases among those infections (Supplementary Table 1). In contrast, reported absolute Access antibiotic usage was less than expected and below 50% of the expected use in Brazil, Mexico, the Philippines, the Russian Federation, and Zambia (observed-to-expected ratio <0.5) in Scenario 1. A similar pattern of Access antibiotic usage was observed among patients aged ≤12 years in most countries except Zambia, where the reported absolute use was above expected.

**Figure 3.**
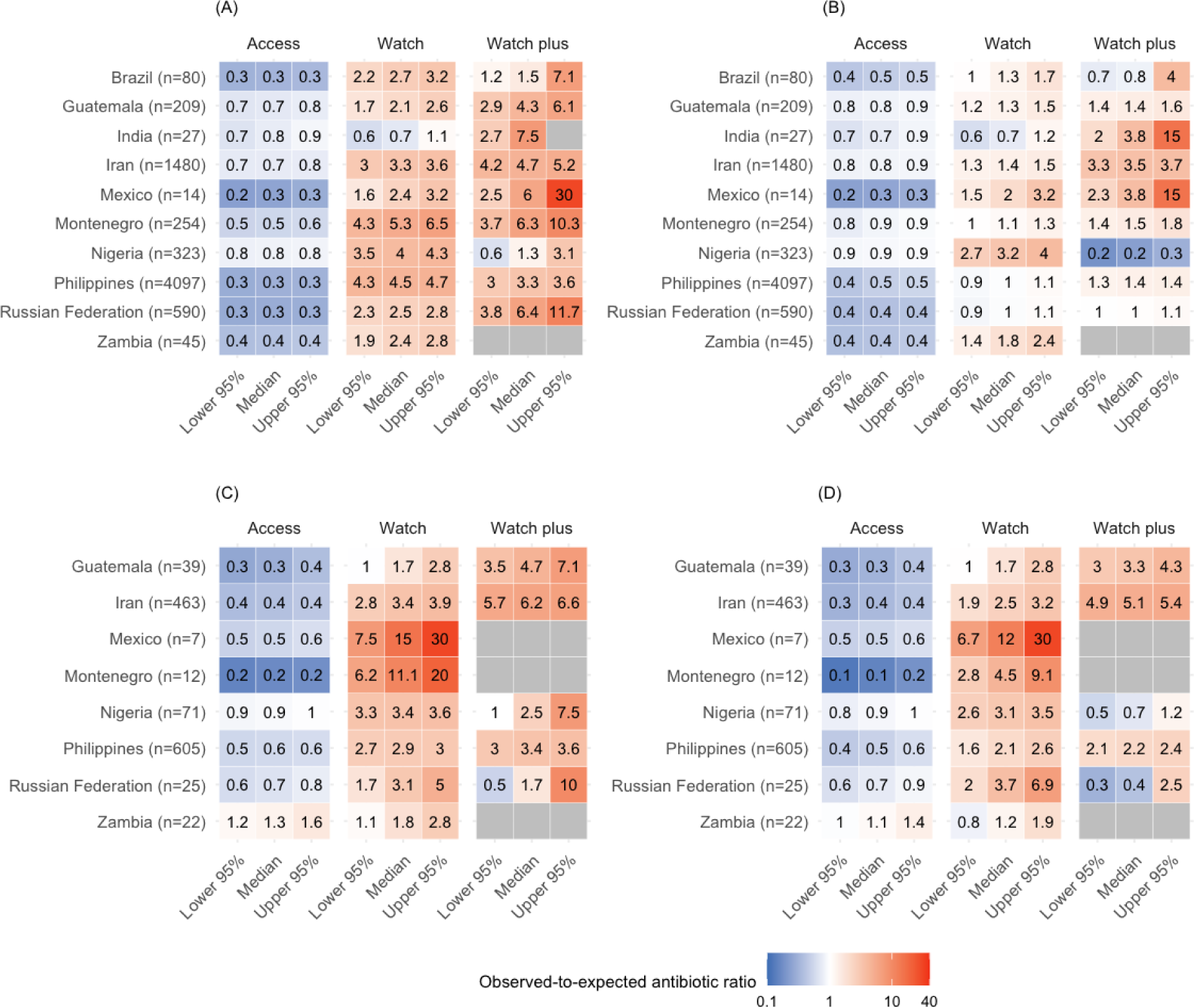
Observed-to-expected ratios of absolute empiric antibiotic use by WHO AWaRe classification across ten countries in 2021. In panel (A) and (B), the observed antibiotic use is crude defined daily dose (DDD) calculated from the Global-PPS data and the expected antibiotic use are estimated DDD from scenario 1 and 2, respectively, for patients aged >12 years. In panel (C) and (D), the observed antibiotic use is crude days of therapy (DOT) calculated from the Global-PPS data and the expected antibiotic use is estimated from scenario 1 and 2, respectively, for patients aged ≤12 years. Values below one indicate lower than expected use, whereas values above one indicate higher than expected use. Grey boxes represent either zero expected use or both zero observed and zero expected use within the indicated AWaRe category.

A similar pattern in both patients aged >12 years and aged ≤12 years was observed in Scenario 2 as in Scenario 1 (Supplementary Figure 24).

### Temporal trend of antibiotic use patterns between 2015 and 2022

We showcased the utility of the observed-to-expected use ratio for monitoring changes in antibiotic use patterns (Figure 4). The ratios were calculated using absolute antibiotic use in DDDs and DOTs for patients aged >12 years and ≤12 years, respectively. The expected antibiotic uses were median estimates of use in DDDs from Scenario 1. There was little evidence of temporal change in the observed-to-expected use ratios for Access and Watch antibiotics across all five countries included in this analysis, with wide credible intervals, in both age groups (Figure 4). The ratios for Access antibiotics fluctuated around 1, indicating concordance between observed and expected absolute antibiotic use. In contrast, the ratios for Watch antibiotics were above 1 across all years, suggesting higher observed than expected use. Among the five countries, Nigeria and the Philippines had the highest observed-to-expected ratios for Watch antibiotics, at around 5, indicating that the observed absolute use was approximately five times higher than expected given the reported local disease burden, and this remained consistent across the study period. There were some indications that the observed-to-expected use ratio for Watch Plus decreased over time and approached the value 1, suggesting increasing alignment between observed and expected absolute antibiotic use among patients aged >12 years (Figure 4A) in the repeated cross-sectional surveys data. These reductions were more pronounced in Brazil, decreasing from an estimated ratio of 9 (observed absolute use was approximately nine times higher than expected) in 2017 to 4 (approximately four times higher) in 2022. This reduction did not coincide with changes in the number of severe cases of infections in Brazil (Figure Supplementary S2). Among patients aged ≤12 years, there was little evidence of changes in the observed-to-expected use of Watch plus antibiotics over time (Figure 4B).

**Figure 4.**
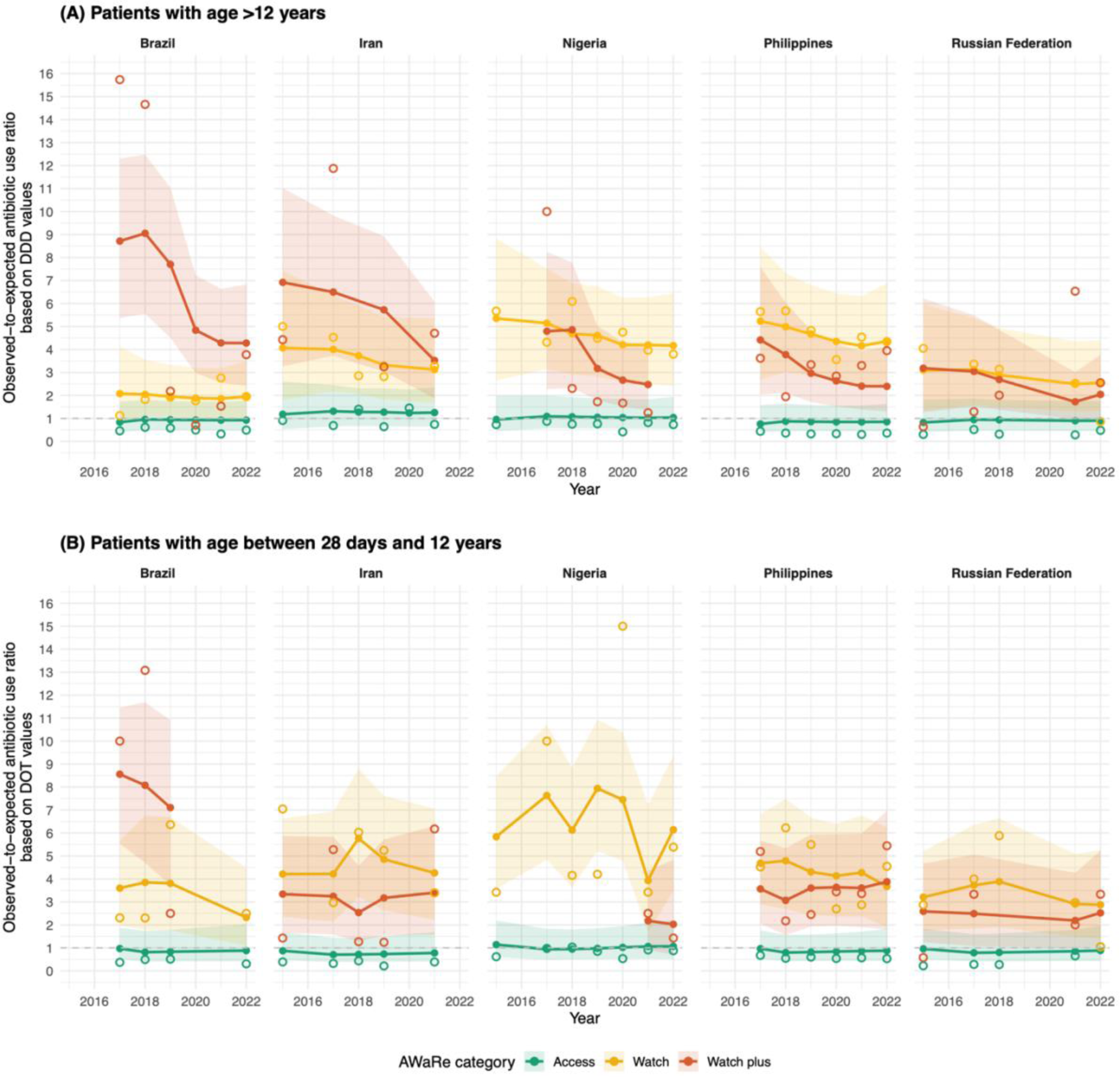
Temporal changes in the observed-to-expected use ratio for the three AWaRe antibiotic groups among patients (A) aged >12 years and (B) 28 days to ≤12 years, based on data from five countries under Scenario 1 between 2015 and 2022. Hollow circles represent the calculated observed-to-expected use ratios, while solid circles represent estimates from a Bayesian multilevel model; shaded areas indicate the 95% credible intervals of the estimates. The horizontal dashed lines mark the value of 1, indicating that observed and expected antibiotic use (measured in DDD and DOT for patients aged >12 years and 28 days to ≤12 years, respectively) are equal.

## Discussion

We developed a benchmarking tool to quantify discrepancies between observed and expected inpatient empiric antibiotic use across ten common infections and surgical prophylaxis, accounting for local infection burden, drug availability, patient characteristics, and guideline-recommended therapy. Applying this framework, we identified consistent underuse of Access antibiotics across multiple indications including skin and soft tissue infections, surgical prophylaxis, hospital-acquired pneumonia, and community-acquired pneumonia in patients aged >12 years, with a substantial overuse of Watch antibiotics. Among these patients, observed Access antibiotic use was 20% to 70% lower than expected. Similarly, among patients aged 28 days to ≤12 years, observed Access antibiotic use was lower than expected in all countries except Zambia. We also observed a modest decline in Watch Plus antibiotic use over time, particularly among patients aged >12 years.

Our study identified substantial discrepancies between observed antibiotic use and that expected under WHO AWaRe book recommendations. These differences could be due to variation in prescribing practices influenced by region-specific pathogen epidemiology.^23^ For example, higher-than-expected use of Watch and Watch Plus antibiotics in community-acquired infections in the Philippines may be driven by empiric use of agents such as ceftriaxone for community-acquired pneumonia based on local clinical guideline, and meropenem for hospital-acquired pneumonia and infections with risk for extended-spectrum beta-lactamase resistance. Furthermore, overuse of Watch antibiotics than optimal was reported more frequent in high-income settings in a recently published modelling study that estimated expected optimal national levels of antibiotics use by AWaRe classification.^24^ Complementing this national-level analysis, our study estimated expected empirical antibiotic use by AWaRe category at the patient-level in hospital settings. Future work could examine microbiological data to identify locally important pathogens for inclusion in treatment guidelines. Notably, our framework can be readily adapted to incorporate such local context, enabling more accurate and policy-relevant assessments of antibiotic use.

Several distinct patterns of antibiotic use were identified from our observed-to-expected ratios heatmap. For instance, in contrast to participating hospitals in other countries, hospitals in Zambia reported a higher proportion of observed empiric Access antibiotic use than expected among patients aged between 28 days and 12 years in 2021. This may be due to a combination of factors, including pressure from caregivers to prescribe antibiotics,^25^ local empirical antibiotic treatment recommendations that differ from those in the WHO AWaRe book, and cost and availability of antibiotics. Our observations from the PPS data are consistent with those of a previous cross-sectional study conducted in a hospital in Zambia, which reported that Access antibiotics were the most commonly prescribed antibiotics and that only four Watch antibiotics (ciprofloxacin, ceftriaxone, cefotaxime, and neomycin) were prescribed during the survey period ^26^. However, the wide uncertainty around the expected antibiotic use estimates for Zambia in our study reflects the limited sample size available for model estimation. Among patients aged >12 years in participating hospitals in Nigeria, we observed lower-than-expected use of empiric Watch Plus antibiotics. However, this finding should not be interpreted as direct evidence of barriers to antibiotic access. Rather, it highlights the need for further investigation into the indications and underlying reasons for this observation, while also emphasising the importance of ensuring adequate access to Watch Plus antibiotics in settings with a high and increasing prevalence of resistant infections.

Priority indications for antimicrobial stewardship varied by country, however bone and joint infections, skin and soft tissue infections, surgical prophylaxis, and community-acquired pneumonia showed higher-than-expected use of Watch antibiotics in most of the included countries across both age groups. Several of these indications, such as surgical prophylaxis and community-acquired pneumonia were also among the most prevalent indications for antibiotic use in our study population. Interventions targeting these indications may therefore have the highest impact in reducing inappropriate use of Watch antibiotics.

Previous studies have applied regression models to estimate expected antibiotic use which was then used to calculate observed-to-expected use ratios.^9–12,14,16^ In contrast, our decision tree approach explicitly incorporates the WHO AWaRe guideline recommendations and drug availability. Thus, our approach closely reflects guideline-based clinical decision-making process underlying empirical antibiotic prescribing and does not require detailed individual-level patient covariates, which are often challenging to obtain in resource and data-limited settings. In addition, our tool generates both relative and absolute measures of expected antibiotic use. Relative measures can inform target setting, evaluation, and monitoring of antibiotic use patterns in hospital settings. Equally important, but often underreported in cross-sectional studies, absolute measures directly quantify discrepancies between observed and expected antibiotic use. Such measures are crucial for drug procurement and financial planning, and can support downstream analyses, including assessments of the cost-effectiveness of antimicrobial stewardship interventions.

Our study has several limitations. First, our estimates of observed antibiotic use were based on Global-PPS data, where participation in the survey was voluntary and the sample may not be nationally representative.^19^ This may limit the generalisability of the findings. Additionally, as point prevalence surveys are conducted on a single day per hospital, our data also do not capture seasonal variation in antibiotic use, and DDDs and DOTs reflect the antibiotic dose or count prescribed on the day of the survey only, not treatment duration. Furthermore, the case-mix observed on the survey day, which is used to estimate expected antibiotic use in our models, may not fully reflect each hospital’s typical disease burden over time. Second, ICU admission and parenteral prescription data were used as proxies in the scenario analyses which may not reflect the actual distribution of severity of cases. These two scenarios were intended to represent lower and upper bounds of case severity, resulting in differences in the estimated use of Watch plus antibiotics (Figure 4), particularly in Mexico and Nigeria. Third, it is possible that the WHO AWaRe book guidance might not be aligned with country-specific empirical treatment guideline for some infections given differences in local epidemiology, AMR patterns and availability of specific antibiotics recommended in the AWaRe book, and therefore these factors are not accounted for in our model.^23^ Fourth, the small sample sizes available for model estimation in several countries including Zambia, Montenegro, India, and Mexico, for model estimation resulted in wide uncertainty intervals around the expected antibiotic use estimates. Moreover, the temporal analysis relied on cross-sectional survey data of different hospitals and assumed that the observed antibiotic use patterns were representative of the country-level patterns over time. Finally, differences in healthcare organisation, infection prevention and control, and healthcare expenditure across countries may influence expected antibiotic use and should be considered when interpreting our findings,

## Conclusion

By identifying specific AWaRe categories and clinical indications where observed use deviates most from expected prescribing patterns, our proposed framework can inform prioritisation of targeted antimicrobial stewardship interventions and guide the development of optimal antibiotic use patient-level targets.

## Data Availability

The Global-PPS datasets used to populate the decision-tree models are available through the Global-PPS project (https://www.global-pps.com/) upon reasonable request and after obtaining all necessary approvals from the respective hospitals.

## Contributors

BSC, MS and CL developed the analysis plan. MMMS built the decision-tree model and populated the model using the aggregated Global-PPS data, WHO-GLASS antimicrobial resistance data, and GRAM data. IP curated and analysed the Global-PPS data. CL and MMMS designed all figures and drafted the first version of the manuscript. All authors revised the manuscript and approved the final version for publication.

## Data sharing

The full analysis code is publicly available at: https://github.com/myomgk/observed_versus_expected_antibiotic_use. The Global-PPS datasets used to populate the decision-tree models are available through the Global-PPS project (https://www.global-pps.com/) upon reasonable request and after obtaining all necessary approvals from the respective hospitals. Other probability parameters used to populate the models are included in the Supplementary table 1.

## Acknowledgement

We thank the members of the Global-PPS network in the participating countries:

Ricardo de Souza Cavalcante (University Hospital - Botucatu Medical School - São Paulo State University, Botucatu, Brazil), Raquel de Oliveira Rabelo (Hospital de Messejana Dr Carlos Alberto Studart Gomes, Fortaleza, Brazil), Francisco Jose Candido da Silva (Hospital Doutor Carlos Alberto Studart Gomes, Fortaleza, Brazil), Evelyne Girão (Walter Cantídio University Hospital of the Federal University of Ceará, Fortaleza, Brazil), Eudes Simões-Neto (Hospital Regional da Baixada Dr Jackson Lago, Federal University of Maranhão - UFMA, Pinheiro, Brazil), Tiago Ferraz (Real Hospital Português de Beneficência em Pernambuco, Recife, Brazil), Júlia Herkenhoff Carijó (Instituto Nacional de Traumatologia e Ortopedia, Rio de Janeiro, Brazil), Filipe Piastrelli (Hospital Alemão Oswaldo Cruz, São Paulo, Brazil), Rosane Coutinho (Hospital Alvorada, São Paulo, Brazil), THAIS GUIMARAES (Hospital do Servidor Público Estadual de São Paulo, São Paulo, Brazil), Brunno Cocentino (Hospital Paulistano, São Paulo, Brazil), Cristhieni Rodrigues (University of São Paulo heart institute and hospital alvorada, São Paulo, Brazil), Maria Luisa Moura (Hospital Vila Nova Star, São Paulo, Brazil), Luciana Passos (Hospital Unimed Vitória, Vitória/Es, Brazil), Randall Lou-Meda (Hospital Roosevelt, Guatemala City, Guatemala), Nancy Sandoval (Hospital Roosevelt, Guatemala City, Guatemala), Brooke M. Ramay (Paul G. Allen School for Global Health, Washington State University Center for Health Studies, Universidad del Valle de Guatemala, Guatemala City, Guatemala), Nuria Chavez (Hospital regional de Zacapa, Zacapa, Guatemala), Olga Adriana Flores Lomán (Hospital Regional de Xalapa Dr. Luis F Nachón, Xalapa, Mexico), Lidija Cizmovic (Institute for Medicines and Medical Devices of Montenegro, Podgorica, Montenegro), Gordana Mijovic (Ministry of Health, Podgorica, Montenegro), Philip Nwajiobi-Princewill (National Hospital Abuja, Abuja, Nigeria), Ubong Udoh (University of Calabar Teaching Hospital, Calabar, Nigeria), Mohammed Manga (Federal Teaching Hospital Gombe Nigeria, Gombe State University, Gombe, Nigeria), Ibinabo Oboro (Rivers State University Teaching Hospital, Port Harcourt, Nigeria), Agantem Ekuma (University of Uyo Teaching Hospital, Uyo, Uyo, Nigeria), Elena Bochanova (Regional Clinical Hospital of Krasnoyarsk, Professor V.F. Voino-Yasenetsky Krasnoyarsk State Medical University, Krasnoyarsk, Russian Federation), Dmitry Popov (A.N. Bakulev National Medical Research Center for Cardiovascular Surgery of the Russian Ministry of Health, Moscow, Russian Federation), Igor Sychev (City Clinical Hospital named after S.S. Yudin; Russian Medical Academy of Continuing Professional Education, Moscow, Russian Federation), Georgiy Melkonyan (Hospital for War veterans #3, Moscow, Russian Federation), Egor Larin (Hospital for War veterans № 3, Moscow, Russian Federation), Ivan Zaharenkov (Moscow City Clinical Hospital named after S.S. Yudin, Moscow, Russian Federation), Oksana Ni (Moscow Multidisciplinary Clinical Center «Kommunarka», Moscow, Russian Federation), Sergey K. Zyryanov (RUDN University named P. Lumumba, Moscow, Russian Federation), Grigorii Sergienko (Kuzbass Children’s Clinical Hospital named after Prof. Yu. E. Malakhovsky, Novokuznetsk, Russian Federation), Elena Elokhina (Regional Clinical Hospital, Omsk, Russian Federation), Nadezhda Zubareva (Perm State Medical University named after Academician E.A. Vagner, Perm, Russian Federation), Vera Vityazeva (Children’s Republican Hospital named after I.N. Grigovicha, Petrozavodsk, Russian Federation), Vladimir Mishchenko (Federal Center of Traumatology and Orthopedics, Smolensk, Russian Federation), Evgeny Dovgan (Smolensk Regional Clinical Hospital, Smolensk, Russian Federation), Roman Kozlov (Smolensk State Medical University, Smolensk, Russian Federation), Anastasia Polyakova (Smolensk State Medical University, Smolensk, Russian Federation), Evgeny Shchetinin (Stavropol State Medical University, Stavropol, Russian Federation), Ulyana Portnyagina (Republican Hospital №2 - Emergency Medical Care Center, North-Easten Federal University, Yakutsk, Russian Federation), Shamil Palyutin (Yaroslavl State Medical University, Yaroslavl, Russian Federation), Lorenzo Bisquera (Asian Hospital And Medical Center, Alabang, Muntinlupa City, The Philippines), Fhebie Zapanta (Corazon Locsin Montelibano Memorial Regional Hospital, Bacolod City, The Philippines), Mary Crist Jamora (Baguio General Hospital and Medical Center, Baguio City, The Philippines), Glory Baltazar (Department of Health, Bataan General Hospital and Balanga City, Balanga, The Philippines), Cristobal Jr. Dumo (St. Luke’s Medical Center, Global City, Bonifacio Global City, Taguig, The Philippines), Jan Steven So (Vicente Sotto Memorial Medical Center, Cebu City, The Philippines), Danice Faith Alombro (Our Lady of The Pillar Medical Center, City of Imus, Cavite, The Philippines), Wynzille Ann De Guzman (Ilocos Training and Regional Medical Center, City of San Fernando, La Union, The Philippines), Vanessa Aira Rivera (Lorma Medical Center, City of San Fernando, La Union, The Philippines), Janelle Morga (Bicol Regional Hospital and Medical Center, Daraga, Albay, The Philippines), Alshan Ruccini Sicat (Jose B.Lingad Memorial General Hospital, Guagua, The Philippines), John Andrew Camposano (Western Visayas Medical Center, Iloilo, The Philippines), Vita Basiao-Gasataya (South Cotabato Provincial Hospital, Koronadal City, South Cotabato, The Philippines), Wynzille Ann De Guzman-Tumbaga (Ilocos Training And Regional Medical Center, La Union, The Philippines), Ma. Theresa Diwata Gagan (Mount Carmel Diocesan General Hospital, Lucena City, Quezon, The Philippines), Gina de Guzman Betito (Makati Medical Center, Makati City, The Philippines), Ma. Tarcela Gler (Makati Medical Center, Makati City, The Philippines), Jay Christian Muere (Makati Medical Center, Makati City, The Philippines), Elfleda Hernandez (Chong Hua Hospital, Mandaue City, Mandaue City, The Philippines), Eva Dizon (RITM, Jose Fabella Memorial Hosp, Manila, The Philippines), Ethel Catherine Antonio (San Lazaro Hospital, Manila, The Philippines), Darel Gresan Quisil (Bukidnon Provincial Hospital Maramag, Maramag, Bukidnon, The Philippines), Shylac Arceo (Research Institute for Tropical Medicine, Metro Manila, The Philippines), Liezel Afaga (Ospital ng Muntinlupa, Muntinlupa City, The Philippines), Desamille Louise Gelle (Asian Hospital and Medical Center, Muntinlupa City, The Philippines), Christine Senete (Asian Hospital And Medical Center, Muntinlupa City, The Philippines), Jemelyn Garcia (Research Institute for Tropical Medicine, Muntinlupa City, The Philippines), Bo Rhenalyn V. (Research Institute for Tropical Medicine, Muntinlupa City, The Philippines), Louella Aquino (James L Gordon Memorial Hospital, Olongapo City, The Philippines), Maryvic Izah Simbajon-Cañedo (Mayor Hilarion A. Ramiro Sr. Medical Center, Ozamiz City, The Philippines), Eileen Kyle Guevara (Adventist Hospital Palawan, Puerto Princesa, The Philippines), Lyza Joy Tadlas (Adventist Hospital Palawan, Puerto Princesa, The Philippines), Sabrina Rei Aguado (De Los Santos Medical Center, Quezon City, The Philippines), Josephine Sengson (Philippine Children’s Medical Center, Quezon City, The Philippines), Kara Marie Venice Lim (Philippine Children’s Medical Center, Quezon City, The Philippines), Jay Ron Padua (Philippine Children’s Medical Center, Quezon City, The Philippines), Melody Tolentino (Philippine Children’s Medical Center, Quezon City, The Philippines), Evan Eriele Dy (Cardinal Santos Medical Center, San Juan City, The Philippines), Stephen Rae Fontanilla (Southern Isabela Medical Center, Santiago, The Philippines), Maizychelle Fami (Eastern Visayas Medical Center, Tacloban City, The Philippines), Graziella May Revilla (Gov. Celestino Gallares Memorial Medical Center, Tagbilaran City, Bohol, The Philippines), Shahreza and CVMC AMS Nadal (Cagayan Valley Medical Center, Tuguegarao City, Cagayan, The Philippines), Sharon D. Bucoy (Zamboanga City Medical Center Pharmacy, Zamboanga City, The Philippines), Kiah Sue Longcob (Zamboanga Doctors’ Hospital, Inc., Zamboanga City, The Philippines), Ellen Marie Tamayo (Tarlac Provincial Hospital, Tarlac City, Philippines), Vivinia Paez (Tarlac Provincial Hospital, Tarlac City, Philippines). Gholamreza Pouladfar (Shiraz University of Medical Sciences, Shiraz, Iran), Mohammad Hassan Aelami (Mashhad University of Medical Sciences, Mashhad, Iran), Sayed Nasser Mostafavi (Isfahan University of Medical Sciences, Isfahan, Iran), Ahmad Shamsizad (Ahvaz (Jundishapur) University of Medical Sciences, Ahvaz, Iran), Ali Hosseininasab (Kerman University of Medical Sciences, Kerman, Iran), Hamid Reza Sherkatolabbasieh (Lorestan University of Medical Sciences, Lorestan, Iran), Negin Nahanmoghaddam (Ardabil University of Medical Sciences, Ardabil, Iran), Zahra Jafarpour (Shiraz University of Medical Sciences, Shiraz, Iran), Ali Amanati (Shiraz University of Medical Sciences, Shiraz, Iran), Mojtaba Shafiekhani (Shiraz University of Medical Sciences, Shiraz, Iran), Behrooz Ataei (Isfahan University of Medical Sciences, Isfahan, Iran), Soodabeh Rostami (Isfahan University of Medical Sciences, Isfahan, Iran), Mehrnaz Aligholipour (Kurdistan University of Medical Sciences, Sanandaj, Iran), Fereshteh Shakornia (Ahvaz (Jundishapur) University of Medical Sciences, Ahvaz, Iran), Mehdi Minaiyan (Isfahan University of Medical Sciences, Isfahan, Iran), Zahra Mehrasa (Lorestan University of Medical Sciences, Lorestan, Iran), Samin Jannatsaray (Faculty of Medicine, Ardabil University of Medical Sciences, Ardabil, Iran).

We thank the members of the ADILA consortium:

Koen B Pouwels (Nuffield Department of Primary Care Health Sciences, University of Oxford, Oxford, UK), Aislinn Cook (Antibiotic Policy Group, School of Health and Medical Sciences, City St George’s, University of London, London, UK), Mike Thorn (Antibiotic Policy Group, School of Health and Medical Sciences, City St George’s, University of London, London, UK), Nam Nguyen (Nuffield Department of Primary Care Health Sciences, University of Oxford, Oxford, UK), Kasim Allel (City St George’s, University of London, UK), Catrin E Moore (Institute for Infection and Immunity, City St George’s, University of London, London, UK)

We thank Gisela Robles Aguilar (Nuffield Department of Medicine, University of Oxford, Oxford, UK) for her inputs on the GRAM data.

## References

1. Goossens H, Ferech M, Vander Stichele R, Elseviers M. Outpatient antibiotic use in Europe and association with resistance: a cross-national database study. Lancet 2005; 365(9459): 579–87.

2. Tang KWK, Millar BC, Moore JE. Antimicrobial Resistance (AMR). British Journal of Biomedical Science 2023; Volume 80 -2023.

3. World Health Organization. AWaRe classification of antibiotics for evaluation and monitoring of use, 2023. 2023. https://www.who.int/publications/i/item/WHO-MHP-HPS-EML-2023.04 (accessed 6 February 2025).

4. Zanichelli V, Sharland M, Cappello B, et al. The WHO AWaRe (Access, Watch, Reserve) antibiotic book and prevention of antimicrobial resistance. Bull World Health Organ 2023; 101(4): 290–6.

5. United Nation General Assembly. Political Declaration of the High-level Meeting on Antimicrobial Resistance. 2024.

6. Mendelson M, Lewnard JA, Sharland M, et al. Ensuring progress on sustainable access to effective antibiotics at the 2024 UN General Assembly: a target-based approach. Lancet 2024; 403(10443): 2551–64.

7. Rhee C, Chen T, Kadri SS, et al. Trends in Empiric Broad-Spectrum Antibiotic Use for Suspected Community-Onset Sepsis in US Hospitals. JAMA Netw Open 2024; 7(6): e2418923.

8. Qu X, Yin C, Sun X, et al. Consumption of antibiotics in Chinese public general tertiary hospitals (2011-2014): Trends, pattern changes and regional differences. PloS one 2018; 13(5): e0196668.

9. Yu KC, Moisan E, Tartof SY, et al. Benchmarking Inpatient Antimicrobial Use: A Comparison of Risk-Adjusted Observed-to-Expected Ratios. Clin Infect Dis 2018; 67(11): 1677–85.

10. Wattier RL, Thurm CW, Parker SK, Banerjee R, Hersh AL. Indirect Standardization as a Case Mix Adjustment Method to Improve Comparison of Children’s Hospitals’ Antimicrobial Use. Clin Infect Dis 2021; 73(5): 925–32.

11. Polk RE, Hohmann SF, Medvedev S, Ibrahim O. Benchmarking risk-adjusted adult antibacterial drug use in 70 US academic medical center hospitals. Clin Infect Dis 2011; 53(11): 1100–10.

12. Khatoun A, Sasaki N, Kunisawa S, Fushimi K, Imanaka Y. Benchmarking broad-spectrum antibiotic use in older adult pneumonia inpatients: a risk-adjusted smoothed observed-to-expected ratio approach. Infect Control Hosp Epidemiol 2025; 46(4): 1–6.

13. Kanerva M, Ollgren J, Lyytikäinen O. Benchmarking antibiotic use in Finnish acute care hospitals using patient case-mix adjustment. J Antimicrob Chemother 2011; 66(11): 2651–4.

14. Cramm SL, Graham DA, Blakely ML, et al. Utility of a Benchmarking Report for Balancing Infection Prevention and Antimicrobial Stewardship in Children With Complicated Appendicitis. Ann Surg 2025; 281(1): 170–5.

15. Fitzpatrick RW, Edwards CM. Evaluation of a tool to benchmark hospital antibiotic prescribing in the United Kingdom. Pharm World Sci 2008; 30(1): 73–8.

16. Sangji NF, Dougherty JM, Tignanelli CJ, et al. Calculation and Feedback of Risk-Adjusted Antibiotic Days as a Process Measure in a Statewide Trauma Collaborative. Am Surg 2024; 90(11): 2814–23.

17. Kamiński B, Jakubczyk M, Szufel P. A framework for sensitivity analysis of decision trees. Central European Journal of Operations Research 2018; 26(1): 135–59.

18. Global PPS. Global PPS Homepage. 2026. https://www.global-pps.com/ (accessed 3 June 2026.

19. Versporten A, Zarb P, Caniaux I, et al. Antimicrobial consumption and resistance in adult hospital inpatients in 53 countries: results of an internet-based global point prevalence survey. The Lancet Global Health 2018; 6(6): e619–e29.

20. WHO Collaborating Centre for Drug Statistics Methodology. ATC/ DDD system https://atcddd.fhi.no/atc_ddd_methodology/purpose_of_the_atc_ddd_system/ (accessed 1 April 2025.

21. World Health Organization. Global Antimicrobial Resistance and Use Surveillance System (GLASS) dashboard. https://worldhealthorg.shinyapps.io/glass-dashboard/_w_12653d9697374014b7c9cab5a8a8e86f/#!/home (accessed 1 April 2025.

22. Institute for Health Metrics and Evaluation. Measuring Infectious Causes and Resistance Outcomes for Burden Estimation (MICROBE) https://vizhub.healthdata.org/microbe/ (accessed 1 April 2025.

23. Chansamouth V, Inlorkham P, Keohavong B, et al. Implementing the WHO AWaRe antibiotic book guidance in lower-resource settings: the case of the Lao PDR. JAC-Antimicrobial Resistance 2024; 6(1).

24. Cook A, Cooper B, Thorn M, et al. Estimating optimal levels of WHO Access, Watch, Reserve (AWaRe) antibiotic use in 186 countries, territories, and areas on the basis of clinical infection and resistance burden. The Lancet Public Health 2026; 11(8): e476–e86.

25. Graham K, Sinyangwe C, Nicholas S, et al. Rational use of antibiotics by community health workers and caregivers for children with suspected pneumonia in Zambia: a cross-sectional mixed methods study. BMC Public Health 2016; 16(1): 897.

26. Mudenda S, Chilimboyi R, Matafwali SK, et al. Hospital prescribing patterns of antibiotics in Zambia using the WHO prescribing indicators post-COVID-19 pandemic: findings and implications. JAC Antimicrob Resist 2024; 6(1): dlae023.

