## Supplementary material for "Quantifying optimal AWaRe antibiotic use patterns in hospitals with a new benchmarking tool applied to cross-sectional point prevalence survey data from 10 countries"

S Figure 18. Proportion of observed versus expected empiric antibiotic use out of the total use within each indication under Scenario 2 in patients > 12 years in 2021 in Brazil, Guatemala, India, Iran and Mexico. 26

S. Figure 19. Proportion of observed versus expected empiric antibiotic use out of the total use within each indication under Scenario 2 in patients > 12 years in 2021in Montenegro, Nigeria, Philippines, Russian Federation and Zambia. ........................................ 27

S Figure 20. Proportion of observed versus expected empiric antibiotic use out of the total use within each indication under Scenario 1 in patients ≤ 12 years in 2021in Guatemala, Iran, Mexico and Montenegro. .28

S Figure 21. Proportion of observed versus expected empiric antibiotic use out of the total use within each indication under Scenario 1 in patients ≤ 12 years in 2021in Nigeria, Philippines, Russian Federation and Zambia. ........................................ 29

S Figure 22. Proportion of observed versus expected empiric antibiotic use out of the total use within each indication under Scenario 2 in patients ≤ 12 years in 2021in Guatemala, Iran, Mexico and Montenegro. .30

S Figure 23. Proportion of observed versus expected empiric antibiotic use out of the total use within each indication under Scenario 2 in patients ≤ 12 years in 2021in Nigeria, Philippines, Russian Federation and Zambia. Purple coloured circular dots indicate the proportion of observed use within each indication......31

S Figure 24. Temporal changes in the observed-to-expected use ratio for the three AWaRe antibiotic groups among patients (A) aged >12 years and (B) 28 days to ≤12 years, based on data from five countries under Scenario 2 between 2015 and 2022. ......................................................................................................32

### Calculation of DDDs and DOTs

We calculated observed antibiotic use in defined daily dose (DDD) for patients >12 years. In brief, prescribed daily doses (PDD) were derived from the reported frequency and unit per dose on the survey day derived from the Global PPS data. Then we divided PDD by the WHO defined DDD, according to the Anatomical Therapeutic Chemical (ATC) classification system (WHO ATC/ DDD) to derive DDD. For patients ≤12 years, one DOT was assigned for each antibiotic administered to a patient per day; thus, a patient receiving two different antibiotics on the same day contributed two DOTs. Expected use was expressed as DDDs in patients aged >12 years and DOTs in patients aged ≤12 years and was calculated using the same approach described above for the observed antibiotic use measures.

### Supplementary tables

S Table 1. Probability parameters to populate the expected use model

| Input variable | Probabilistic distribution | Source of data and justification |
|---|---|---|
| **Both age groups** | | |
| Probability of prescribing first-choice antibiotics | beta(8, 2)<br>Range: 0.32 - 0.99<br>IQR: 0.72 - 0.89 | Assumed parameter as all the recommended first-choice antibiotics in the WHO AWaRe book are listed in the essential medicine lists (EMLs) and they are expected to be available. Thus, we assume the probability of being prescribed a first-choice antibiotics is around 80%. |
| Probability of severe disease in patients with community acquired pneumonia, intra-abdominal infection, and acute pyelonephritis | **Scenario 1**<br>beta($1 + \alpha_1$, $1 + \beta_1$)<br><br>**Scenario 2**<br>beta($1 + \alpha_2$, $1 + \beta_2$) | **Scenario 1**<br>$\alpha_1$ = patients admitted to ICU<br>$\beta_1$ = patients not admitted to ICU<br><br>**Scenario 2**<br>$\alpha_2$ = patients who received at least one parental antibiotic<br>$\beta_2$ = patients who received only oral antibiotics<br><br>Given the unavailability of empirical data on patient severity, we assumed patients admitted to ICU to be severe cases in scenario 1 and patients who received at least one parental antibiotic to be severe cases in scenario 2. |
| Probability of high-risk multi-drug resistant (MDR) infection in patients with hospital acquired pneumonia (non-VAP) | beta(651, 1098)<br>Range: 0.33 - 0.42<br>IQR: 0.36 - 0.38 | The data came from a 10-year prospective observational study in China on clinical and microbiological characteristics of adults with hospital acquired pneumonia. The study defined MDR as isolates resistant to three or more different antimicrobial classes, and all methicillin-resistant *S. aureus* (MRSA).<br>(doi: 10.1007/s10096-020-04046-9) |
| Probability of necrotizing fasciitis (NF) in patients with skin and soft tissue infection (SSTI) | beta(9.01, 291.4)<br>Range: 0.005 - 0.074<br>IQR: 0.02 - 0.04 | NF is a life-threatening, but rare infection. Literature suggests NF constitutes less than 5% of overall SSTI infections. Thus, we estimated beta probability distribution parameters assuming an estimate of 3% (95% CI: 1 – 5%).<br>doi:10.1371/journal.pntd.0008887,<br>doi: 10.1016/j.amsu.2020.09.013 |
| Probability of *Streptococcus pyogenes* (Group A Streptococcus) infection in patients with necrotizing fasciitis | beta(132, 166)<br>Range: 0.35 - 0.53<br>IQR: 0.42 - 0.46 | A systematic review of necrotising fasciitis in children from its first description in 1930 to 2018 (doi: 10.1186/s12879-019-3941-3) |

| | | |
|---|---|---|
| Probability of suspected/ high risk setting for ESBL infection in severe cases of intra-abdominal infection, acute pyelonephritis, febrile neutropenia, and sepsis. | beta($\alpha$, $\beta$) | Country- and year-specific ESBL prevalence data (bloodstream bacteriologically confirmed infections) from the WHO GLASS dashboard.<br>$\alpha$ = (proportion of *Escherichia coli* isolates resistant to third-generation cephalosporin) × total number of hospitalised patients on empirical antibiotics (country- and year-specific data from the Global-PPS).<br>$\beta$ = (1- proportion of *E. coli* isolates resistant to third-generation cephalosporin) × total number of hospitalised patients on empirical antibiotics (country- and year-specific data from the Global-PPS).<br>For years with fewer than 30 interpretable isolates data from the most recent previous year was used. The WHO GLASS AMR data was not available for Montenegro, Russian Federation, Mexico and Guatemala. For these countries, (unpublished) data from the Global Research on Antimicrobial Resistance (GRAM) study was used. The high risk setting for ESBL infection is defined if the prevalence exceeds 20%. |
| Probability of suspected/ high risk of resistant gram-negative bacterial infection in febrile neutropenia | Same as above | Same as above |
| Probability of suspected MRSA infection in severe cases of skin and soft tissue infection, bone and joint infections, and febrile neutropenia | beta($\alpha$, $\beta$) | Country- and year-specific MRSA prevalence data (bloodstream bacteriologically confirmed infections) from the WHO GLASS dashboard.<br>$\alpha$ = (proportion of *S. aureus* isolates resistant to methicillin) × total number of hospitalised patients on empirical antibiotics (country- and year-specific data from the Global-PPS).<br>$\beta$ = (1- proportion of *S. aureus* isolates resistant to methicillin) × total number of hospitalised patients on empirical antibiotics (country- and year-specific data from the Global-PPS).<br>For years with interpretable isolates less than 30, data from the most recent previous year was used. The WHO GLASS AMR data was not available for |

| | | |
|---|---|---|
| | | Montenegro, Russian Federation, Mexico and Guatemala. For these countries, (unpublished) data from the Global Research on Antimicrobial Resistance (GRAM) study was used. |
| **Patients aged >12 years** | | |
| Probability of underlying causes of sepsis (unknown origin, enteric fever, meningitis, lower respiratory tract infection, intra-abdominal infection, skin and soft tissue infection, acute pyelonephritis) | dirichlet(0.752, 0.055, 0.016, 0.128, 0.017, 0.008, 0.024) x total number of hospitalised patients on empirical antibiotics (country- and year-specific data from the Global-PPS) | The dirichlet probability distribution parameters were referenced from the Global, regional, and national sepsis incidence and mortality, 1990-2017: analysis for the Global Burden of Disease Study.<br>DOI: 10.1016/S0140-6736(19)32989-7 |
| **Patients aged ≤ 12 years** | | |
| Probability of treatment failure (no response to ampicillin + gentamicin) after 48 hours | beta(5.4, 12.6)<br>Range: 0.07 - 0.72<br>IQR: 0.22 - 0.37 | Literature suggests a wide range of treatment failure to first-line treatment (penicillin/ ampicillin + gentamicin). Thus, we estimated beta probability distribution parameters assuming an estimate of 30% (95% CI: 8 – 50.4%).<br>doi: 10.1093/cid/cis1201,<br>doi:10.1371/journal.pone.0278938,<br>doi:10.1371/journal.pone.0122052.<br>doi:10.1371/journal.pone.0281209. |
| Probability of HIV infection in patients ≤ 12 years with severe community acquired pneumonia | <u>African countries</u><br>beta(218, 1097)<br>Range: 0.13 - 0.20<br>IQR: 0.16 - 0.17<br><br><u>Non-African countries</u><br>beta(33, 2247)<br>Range: 0.008 - 0.026<br>IQR: 0.013 - 0.016 | The data comes from screening data of the study “Causes of severe pneumonia requiring hospital admission in children without HIV infection from Africa and Asia: the PERCH multi-country case-control study”.<br>Ref: 10.1016/S0140-6736(19)30721-4 (PERCH study) |

### Supplementary figures

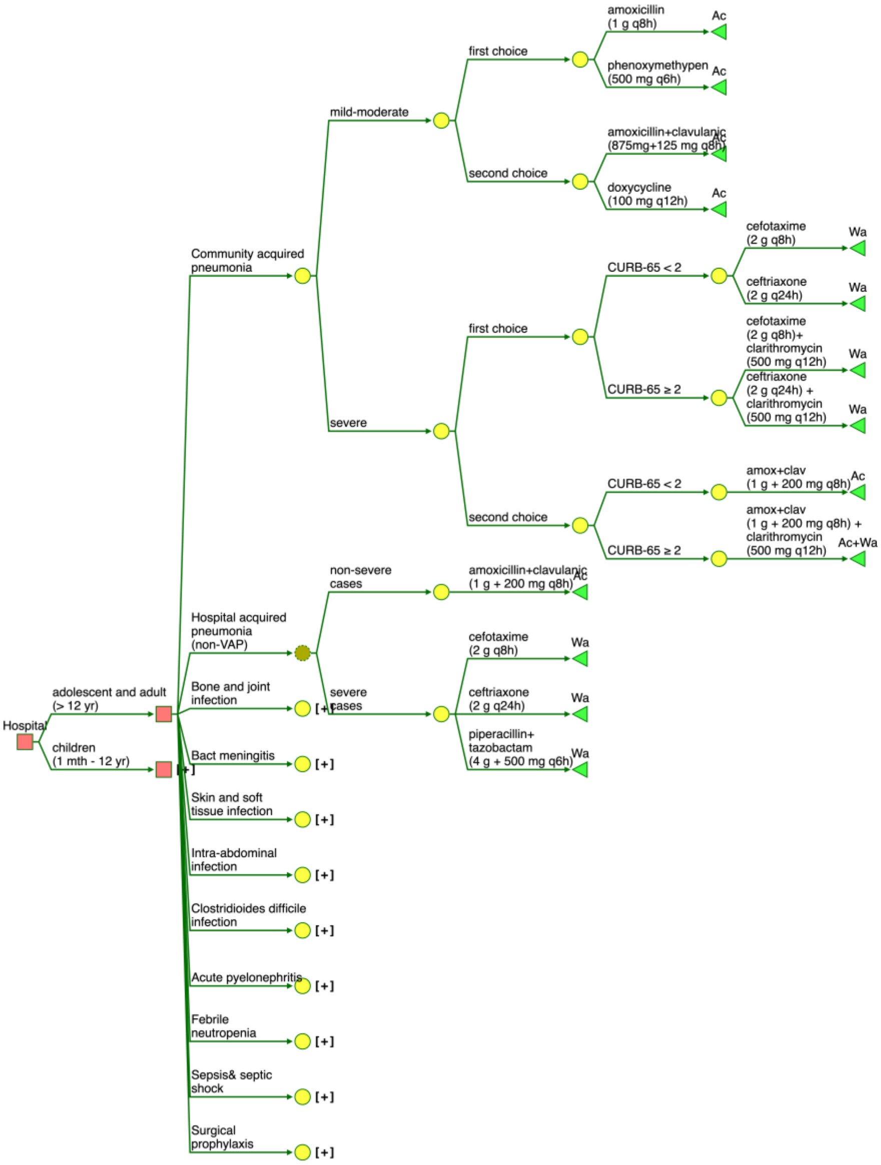

S Figure 1. Decision tree model showing empirical treatment of community acquired pneumonia (CAP) and hospital acquired pneumonia (HAP) for patients aged > 12 years based on the WHO AWaRe book guidelines. (Ac = Access, Wa = Watch, WaP = Watch plus)

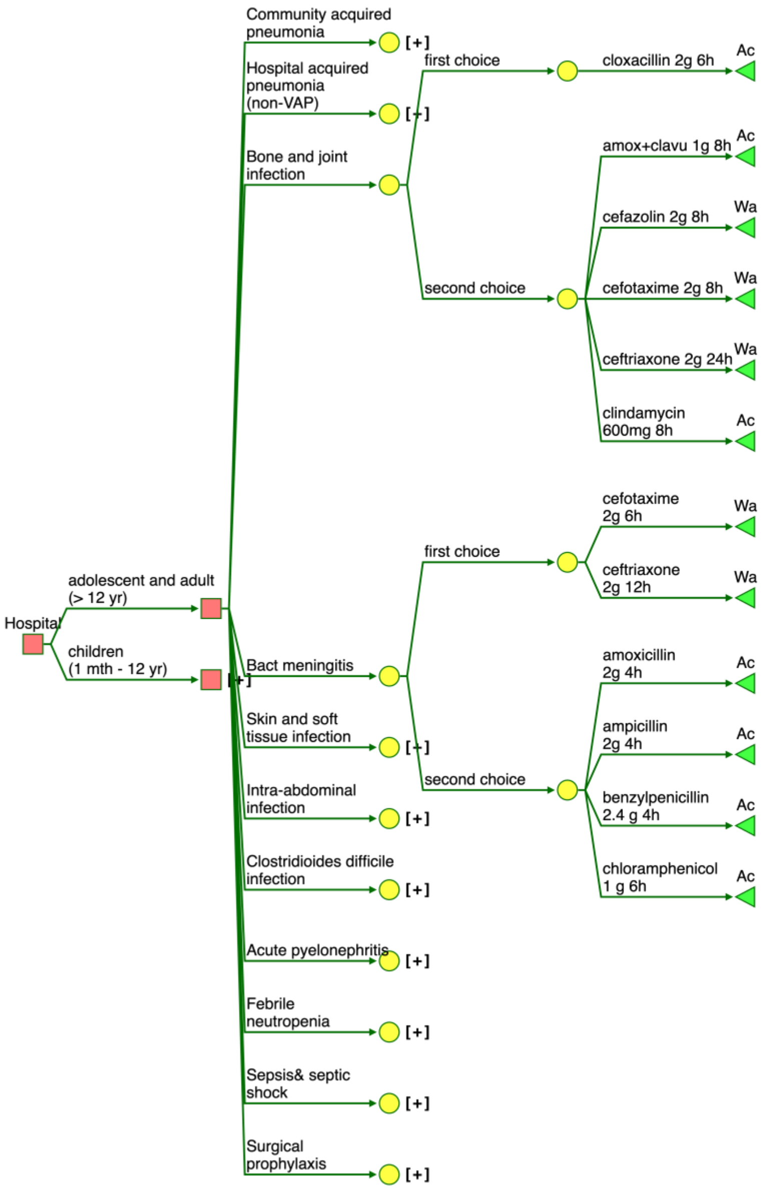

S Figure 2. Decision tree model showing empirical treatment of bone and joint infection, and bacterial meningitis for patients aged > 12 years based on the WHO AWaRe book guidelines. (Ac = Access, Wa = Watch, WaP = Watch plus)

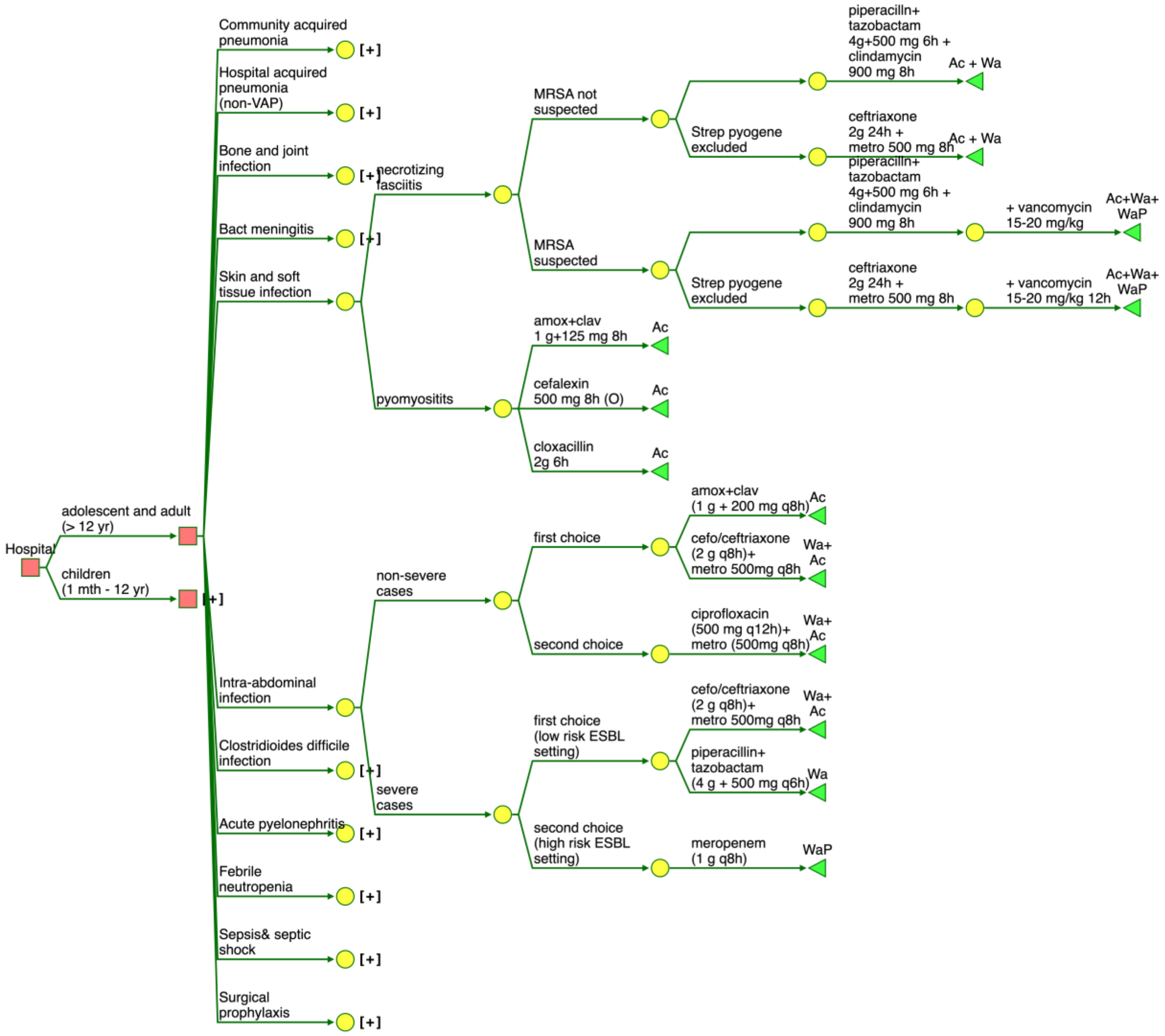

S Figure 3. Decision tree model showing empirical treatment of skin and soft-tissue infection, and intra-abdominal infection for patients aged > 12 years based on the WHO AWaRe book guidelines. (Ac = Access, Wa = Watch, WaP = Watch plus)

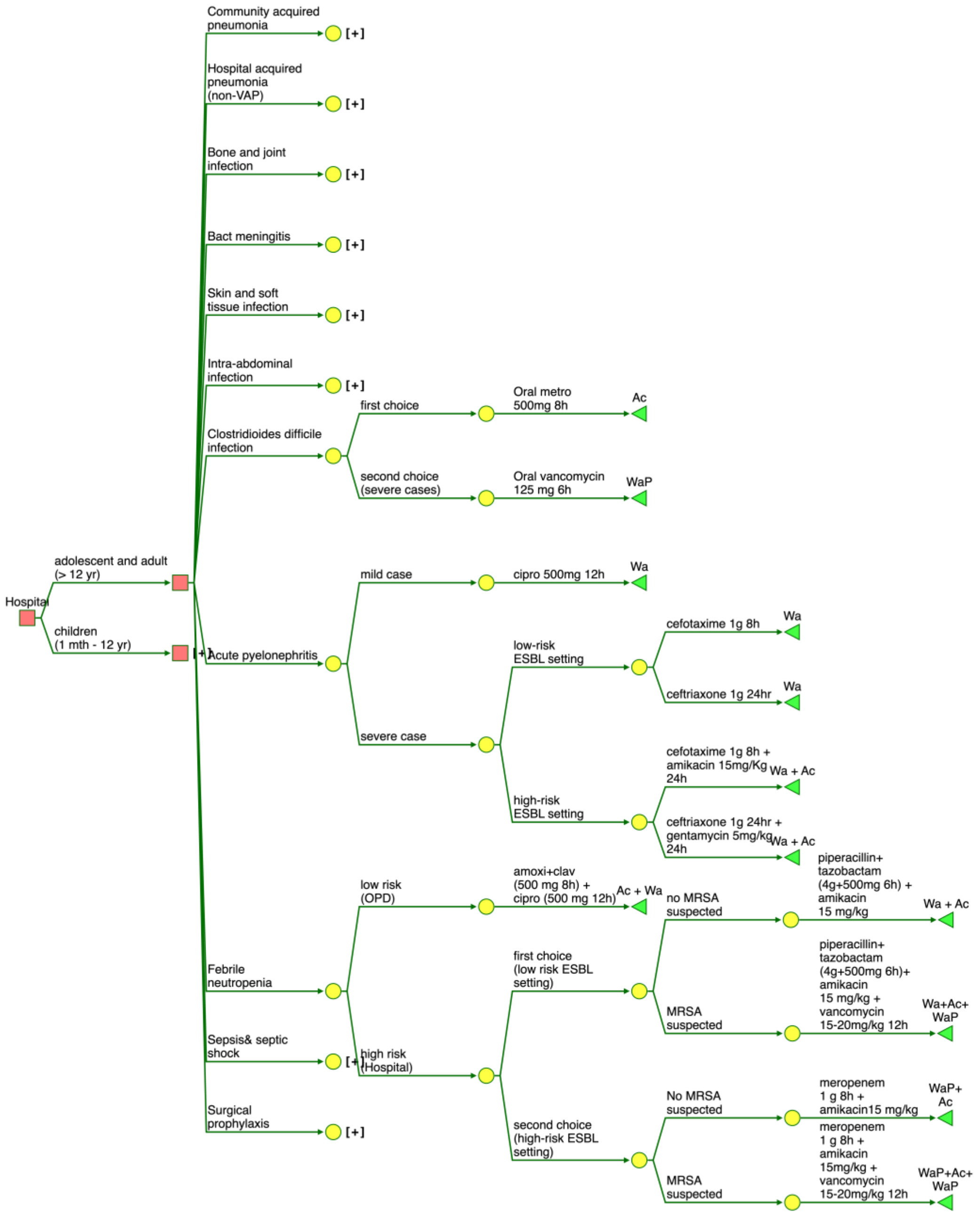

S Figure 4. Decision tree model showing empirical treatment of *Clostridioides difficile* infection, acute pyelonephritis and febrile neutropenia for patients aged > 12 years based on the WHO AWaRe book guidelines. (Ac = Access, Wa = Watch, WaP = Watch plus)

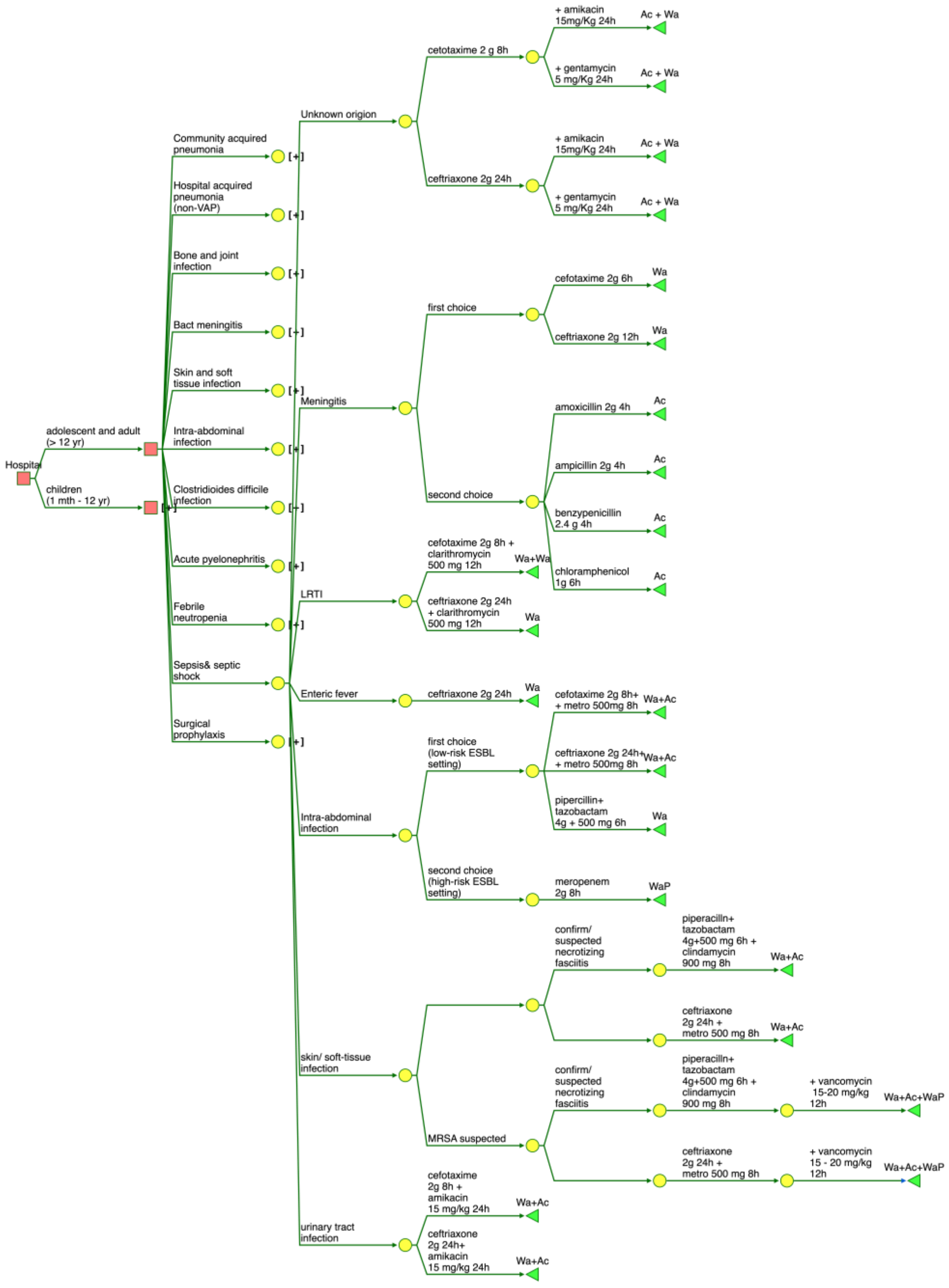

S Figure 5. Decision tree model showing empirical treatment of sepsis and septic shock in patients aged > 12 years based on the WHO AWaRe book guidelines. (Ac = Access, Wa = Watch, WaP = Watch plus)

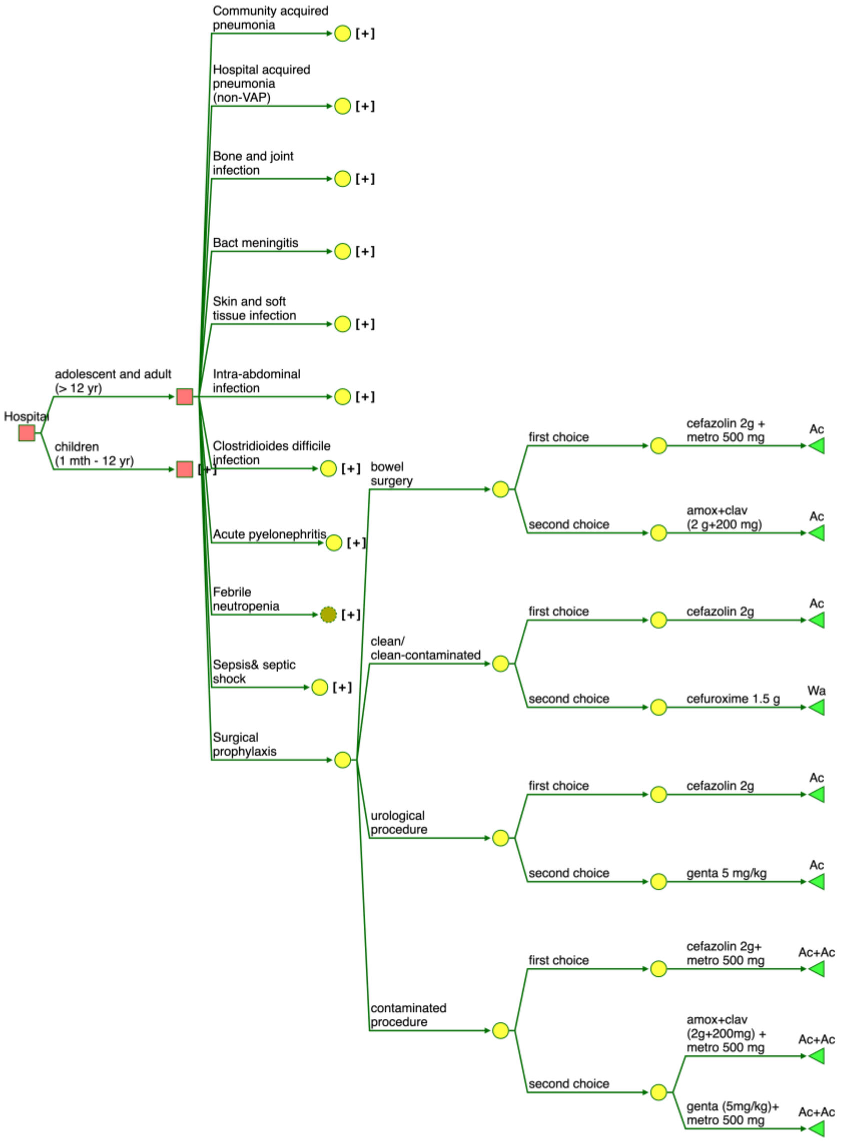

S Figure 6. Decision tree showing empirical antibiotic recommendation for surgical prophylaxis in patients aged > 12 years based on the WHO AWaRe book guidelines. (Ac = Access, Wa = Watch, WaP = Watch plus)

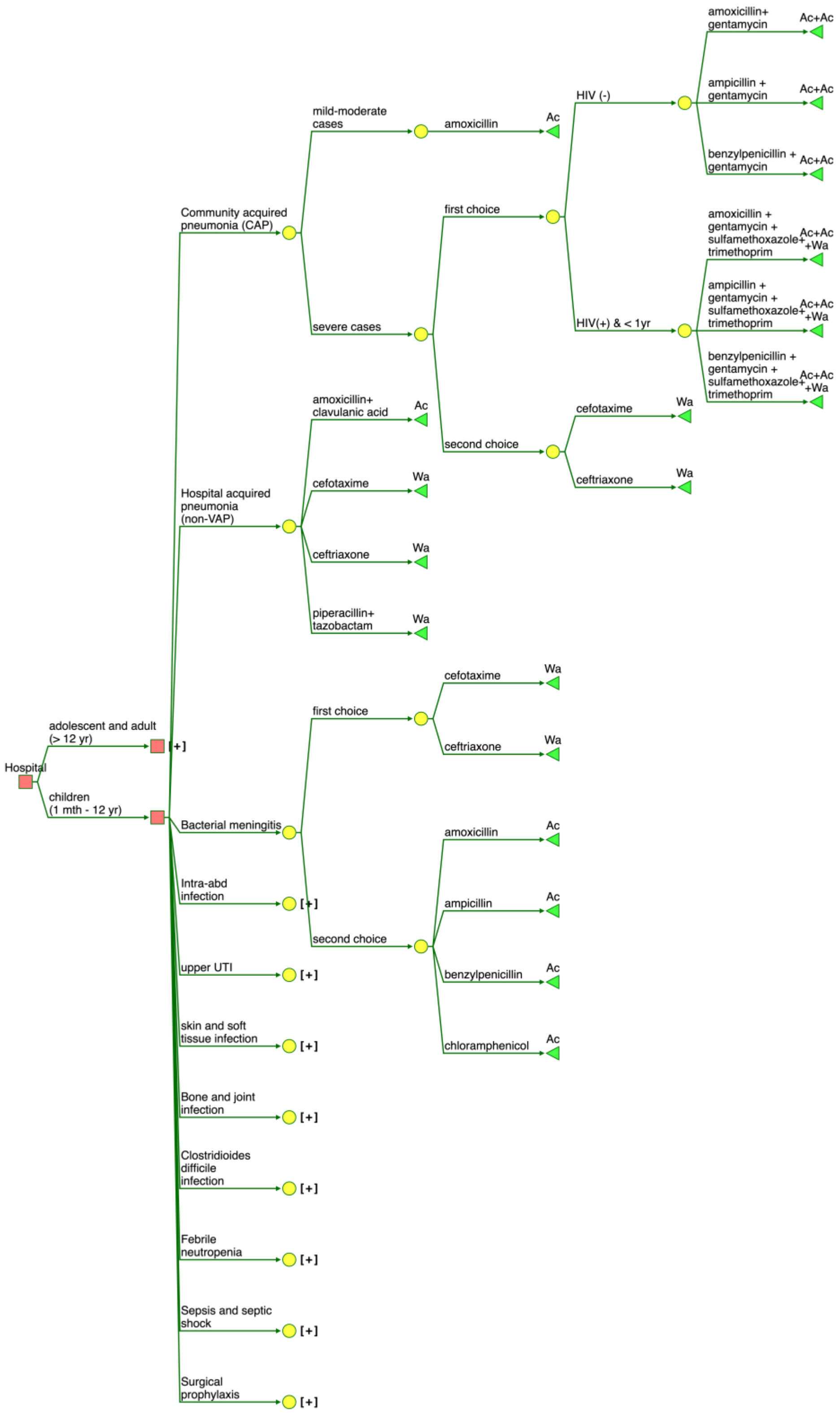

S Figure 7. Decision tree model showing empirical treatment of community acquired pneumonia, non-ventilator associated hospital acquired pneumonia and bacterial meningitis in patients aged ≤ 12 years based on the WHO AWaRe book guidelines. (Ac = Access, Wa = Watch, WaP = Watch plus)

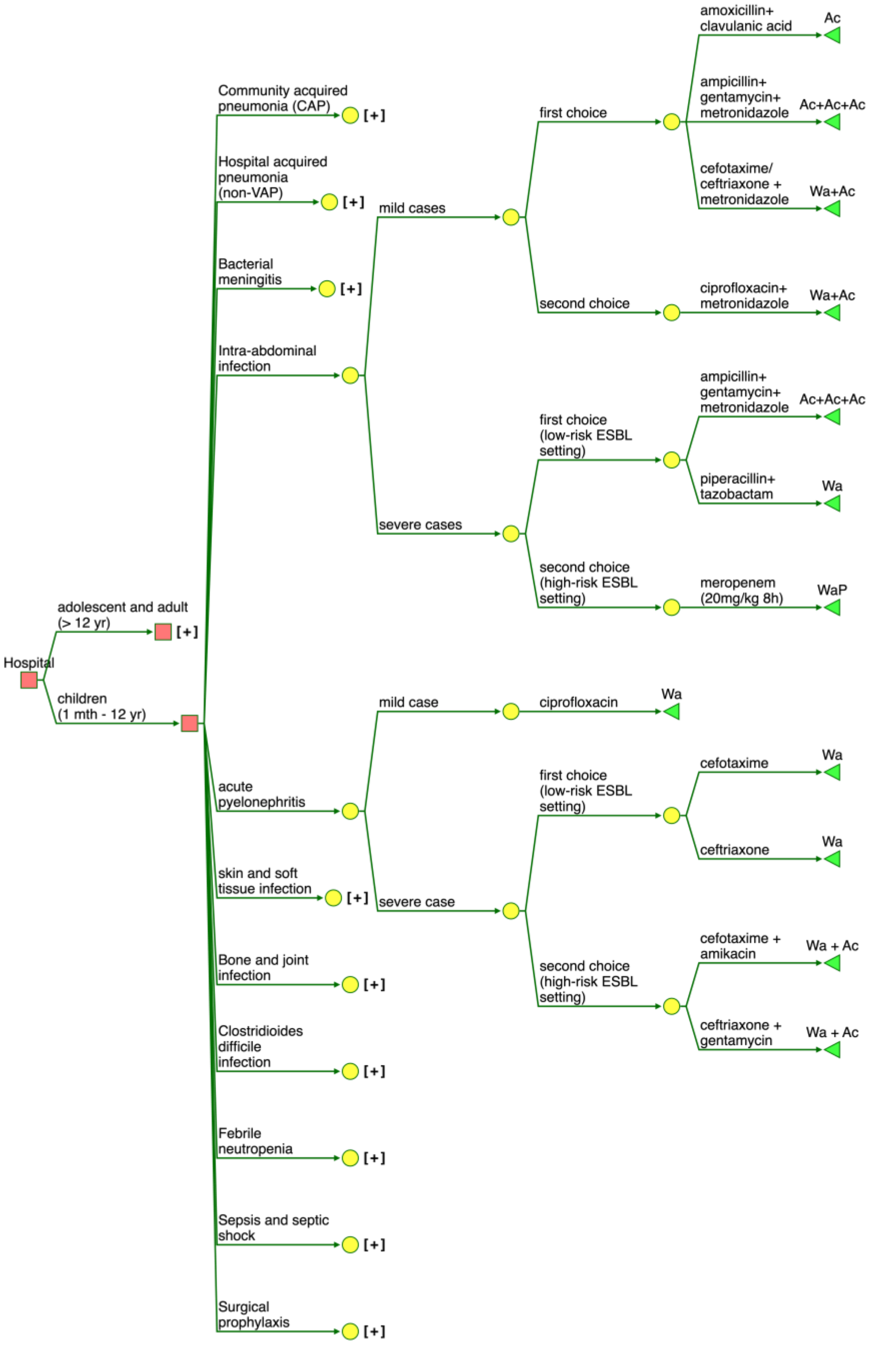

S Figure 8. Decision tree model showing empirical treatment of intra-abdominal infection and acute pyelonephritis in patients aged ≤ 12 years based on the WHO AWaRe book guidelines. (Ac = Access, Wa = Watch, WaP = Watch plus)

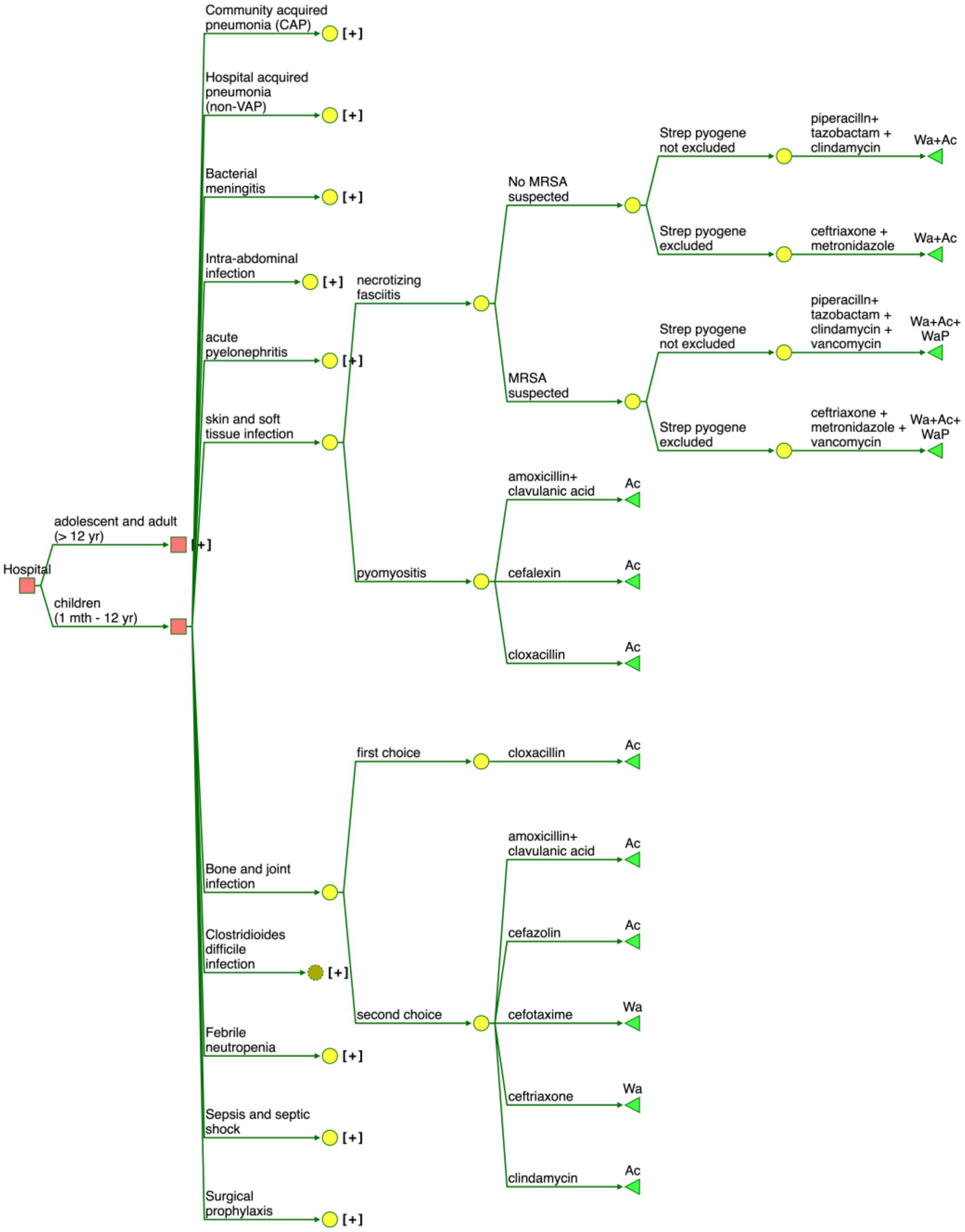

S Figure 9. Decision tree model showing empirical treatment of skin and soft-tissue infection and bone and joint infection in patients aged ≤ 12 years based on the WHO AWaRe book guidelines. (Ac = Access, Wa = Watch, WaP = Watch plus)

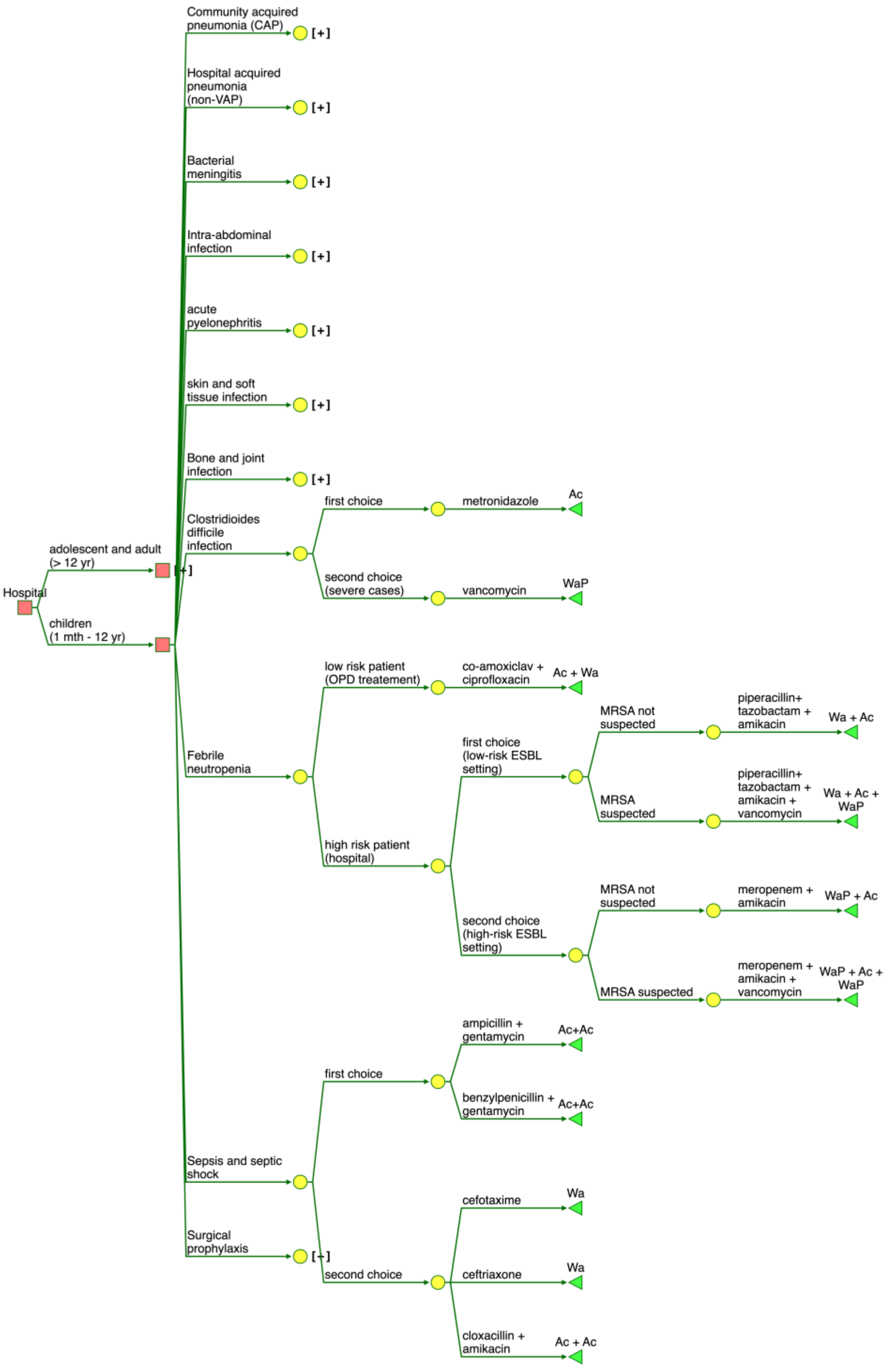

S Figure 10. Decision tree model showing empirical treatment of *Clostridioides difficile* infection, febrile neutropenia and sepsis and septic shock in patients aged ≤ 12 years based on the WHO AWaRe book guidelines. (Ac = Access, Wa = Watch, WaP = Watch plus)

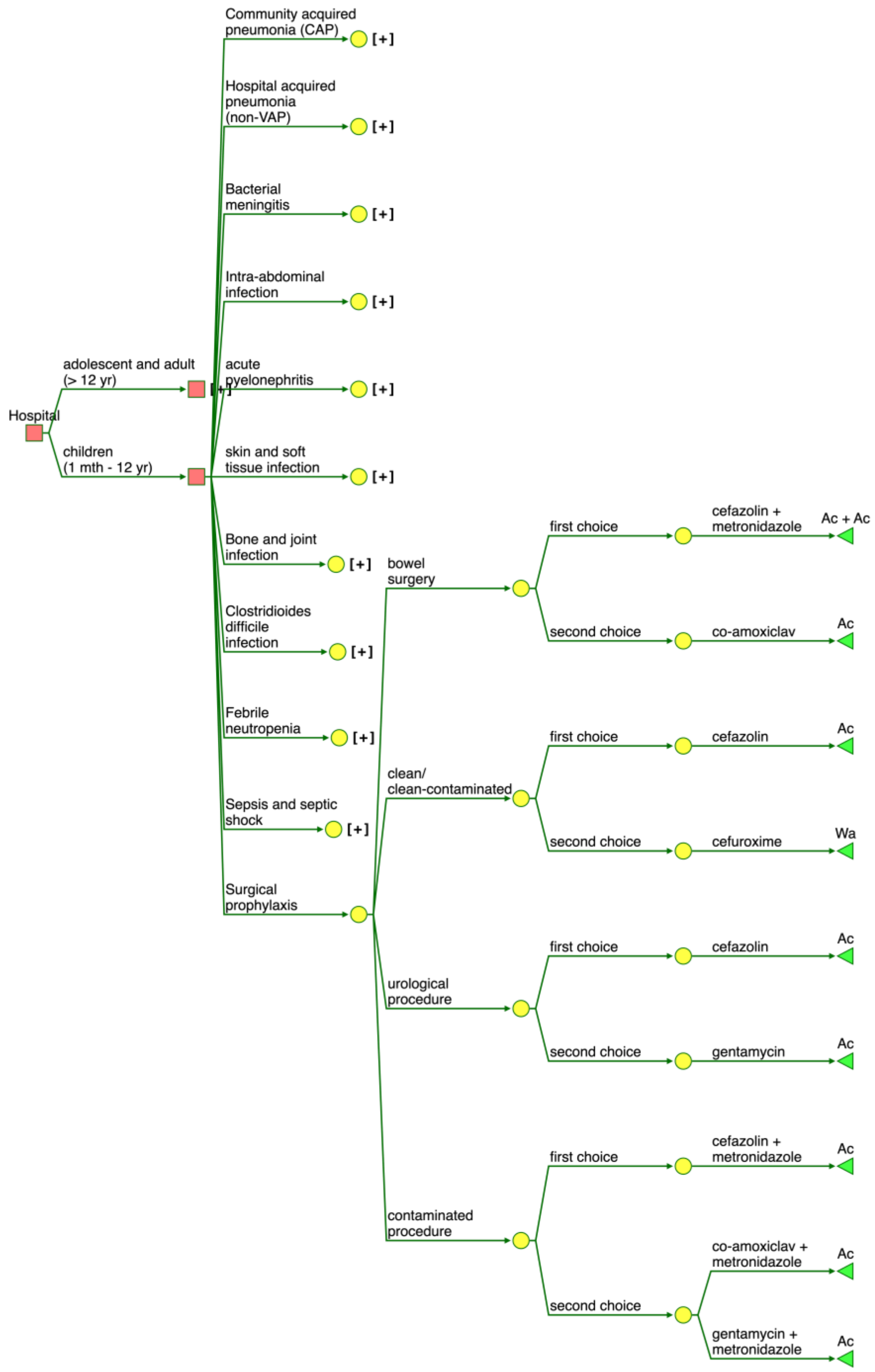

S Figure 11. Decision tree showing empirical antibiotic recommendation for surgical prophylaxis in patients aged ≤ 12 years based on the WHO AWaRe book guidelines. (Ac = Access, Wa = Watch, WaP = Watch plus)

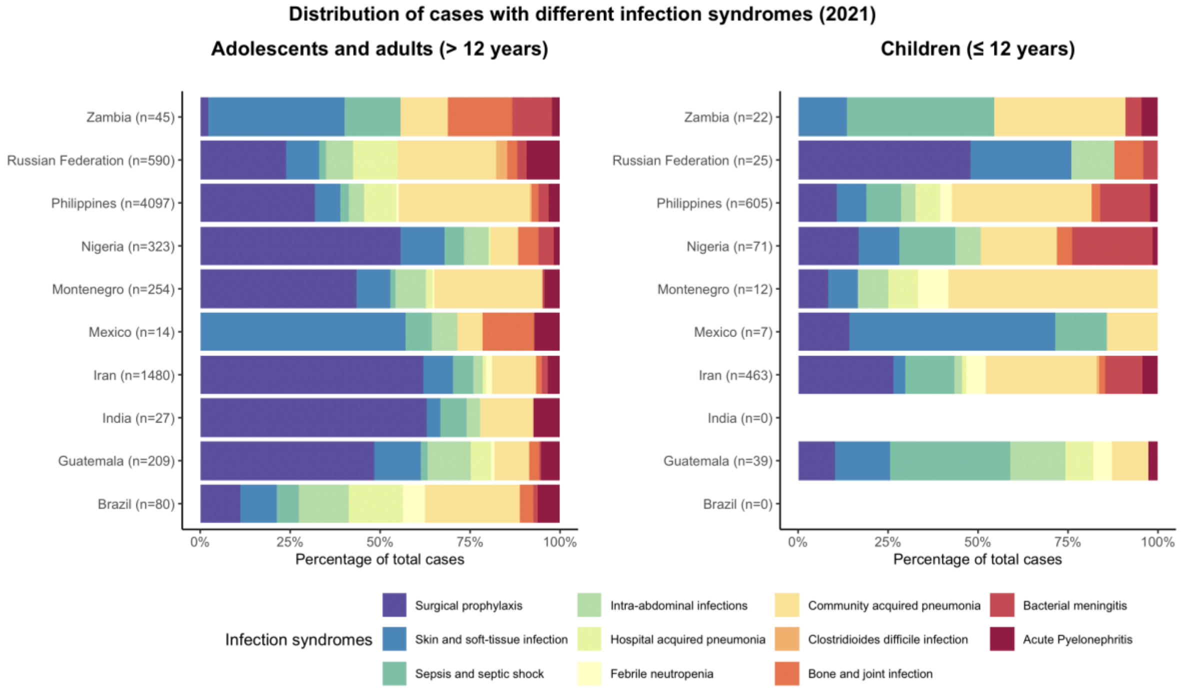

S Figure 12. Distribution of infection syndromes (indications) in 2021. The country labels include the total number of patients across all years.

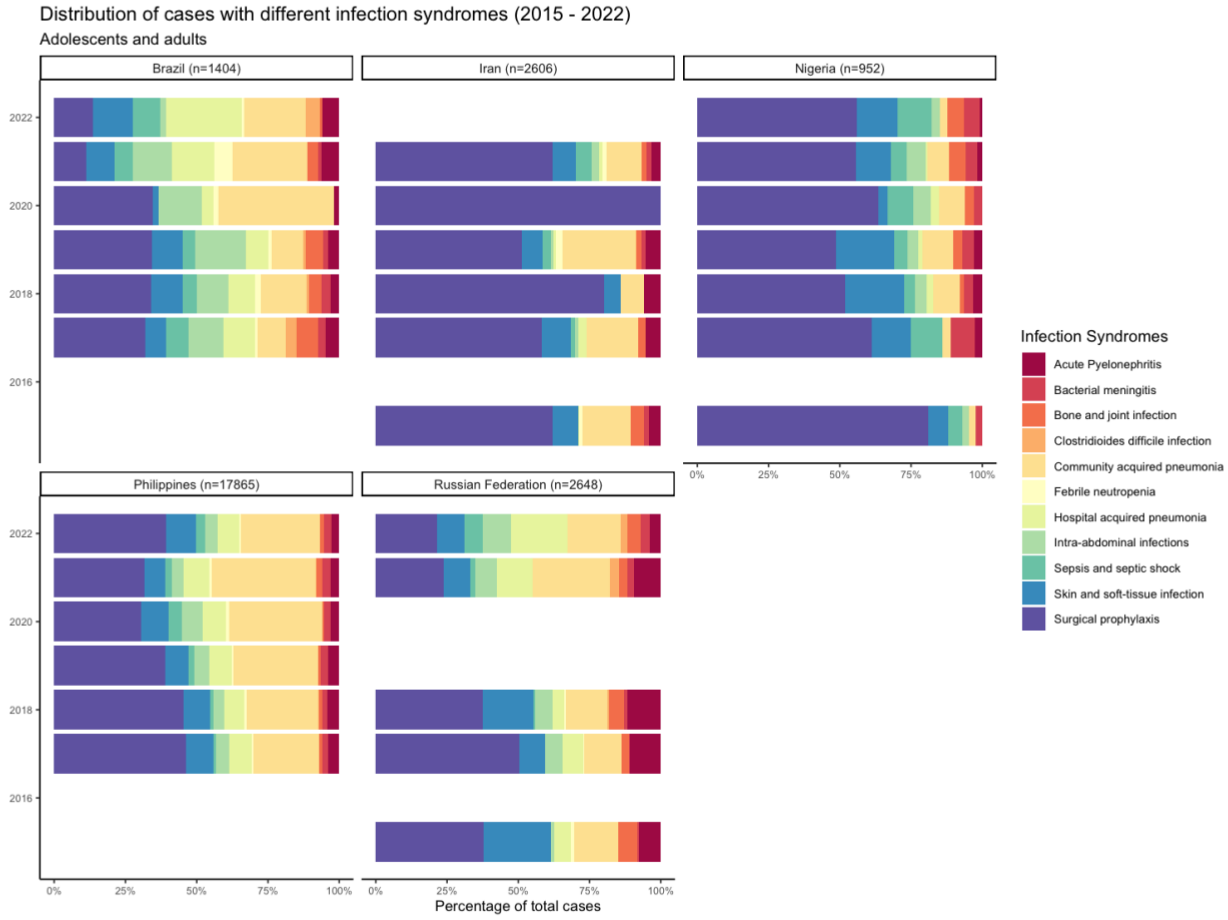

S Figure 13. Distribution of infection syndromes (indications) in patients > 12 years across five countries from 2015 to 2022. The country labels include the total number of patients across all years.

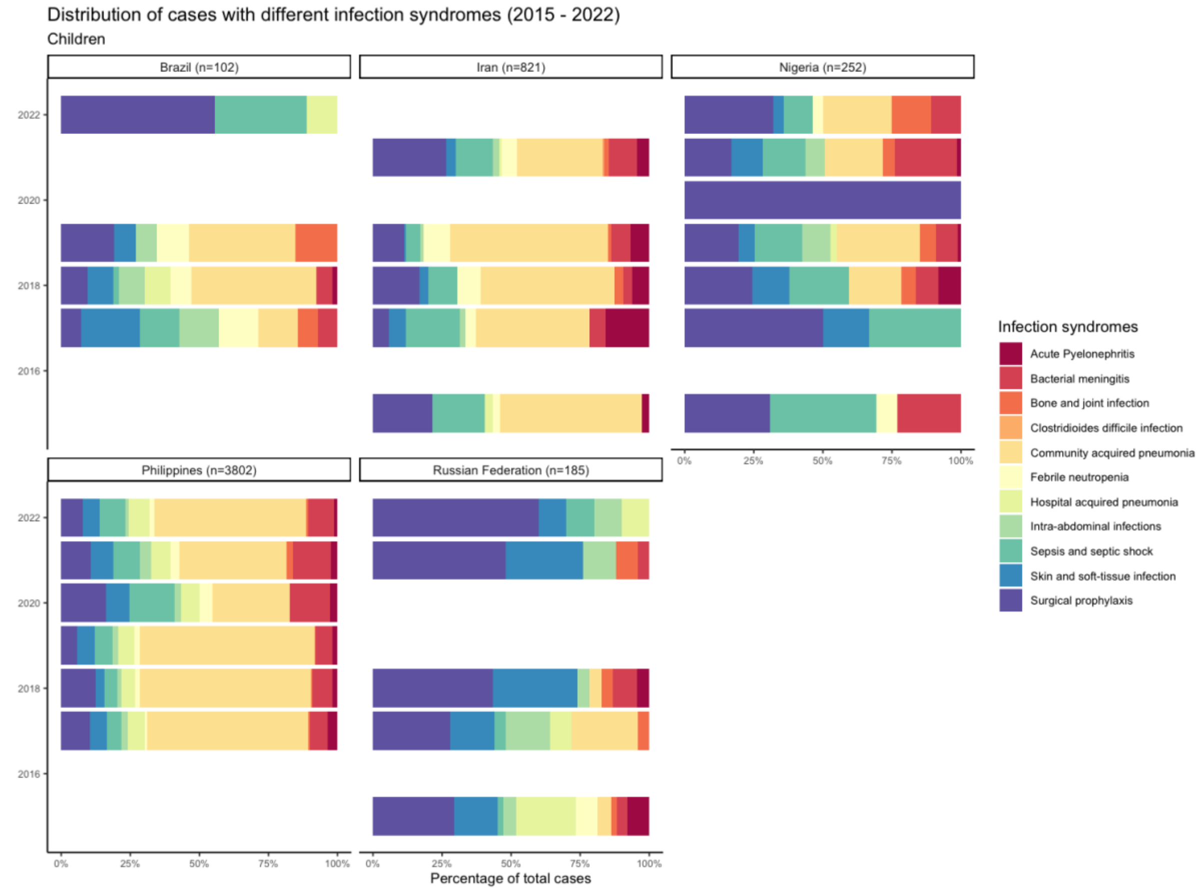

S Figure 14. Distribution of infection syndromes (indications) in patients ≤ 12 years across five countries from 2015 to 2022. The country labels include the total number of patients across all years.

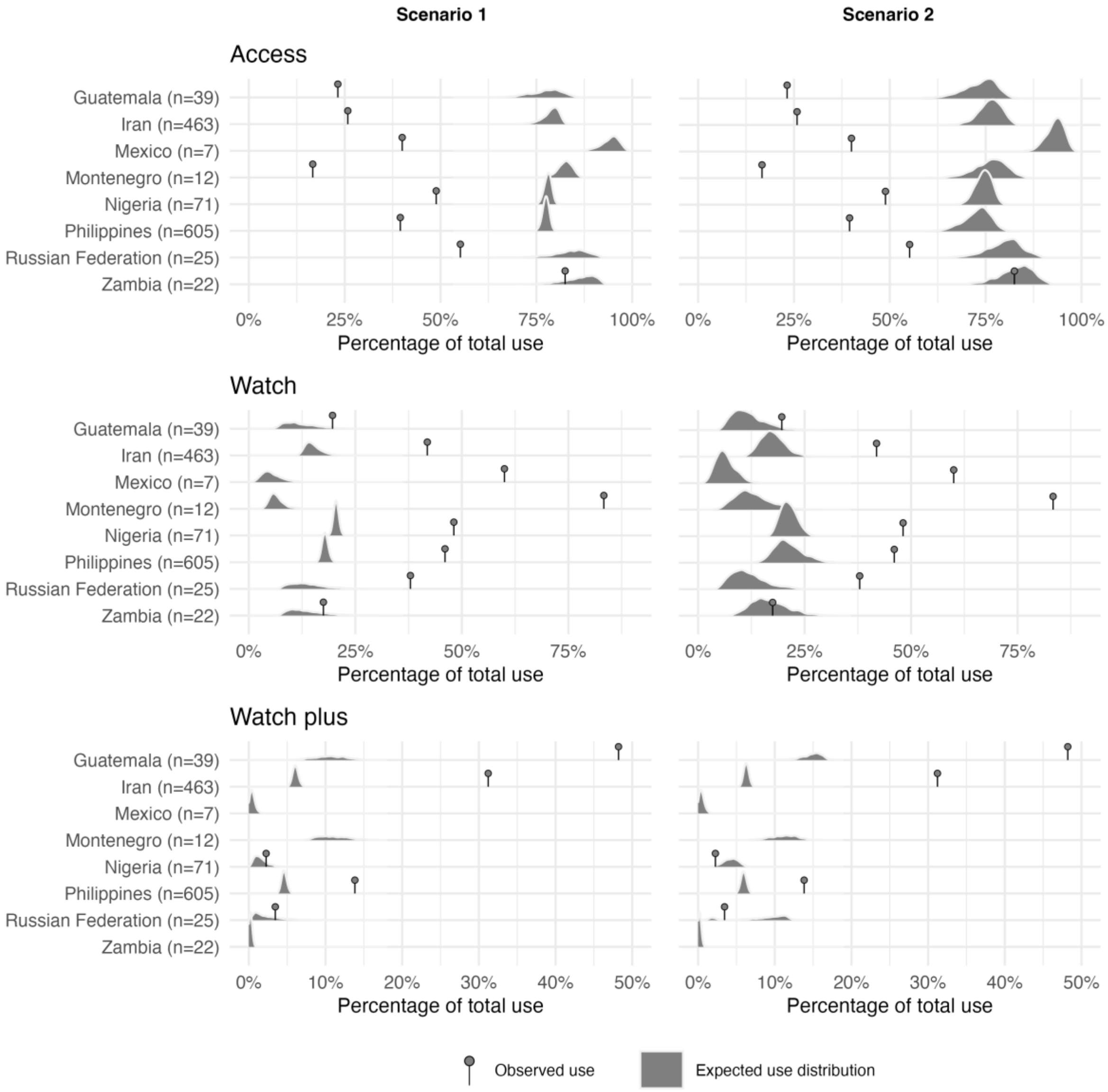

S Figure 15. The proportion of observed versus expected antibiotic use relative to total antibiotic use across the three AWaRe categories in patients ≤ 12 years. The lollipop markers indicate observed antibiotic use, and the density plots represent expected distribution of antibiotic use, expressed as a percentage of total day of therapy (DOT) among patients ≤ 12 years on the day of the point prevalence survey in 2021. Data unavailable for Brazil and India.

**Proportion of observed versus expected antibiotic use in patients ≤ 12 years (2021)**
Access antibiotics comprised the largest share of expected use in patients ≤ 12 years in all countries, followed by Watch and Watch-plus antibiotics (S Figure 15). Under scenario 1, Access antibiotics were expected to account for at least 70% of total use across all countries, while expected use of Watch and Watch-plus antibiotics was comparatively lower, remaining below 25% and 10%, respectively. In contrast, observed proportions deviated markedly from the expected, mirroring patterns seen among patients >12 years. Access antibiotics accounted for a smaller share of total use than expected, whereas Watch antibiotics accounted for a larger share in all countries except Zambia, where observed proportions fell within the expected range. In scenario 2, the gap between observed and expected proportions of Access and Watch antibiotics narrowed compared with scenario 1, consistent with patterns observed among patients > 12 years. No observed use of Watch plus antibiotics was reported in Mexico and Zambia.

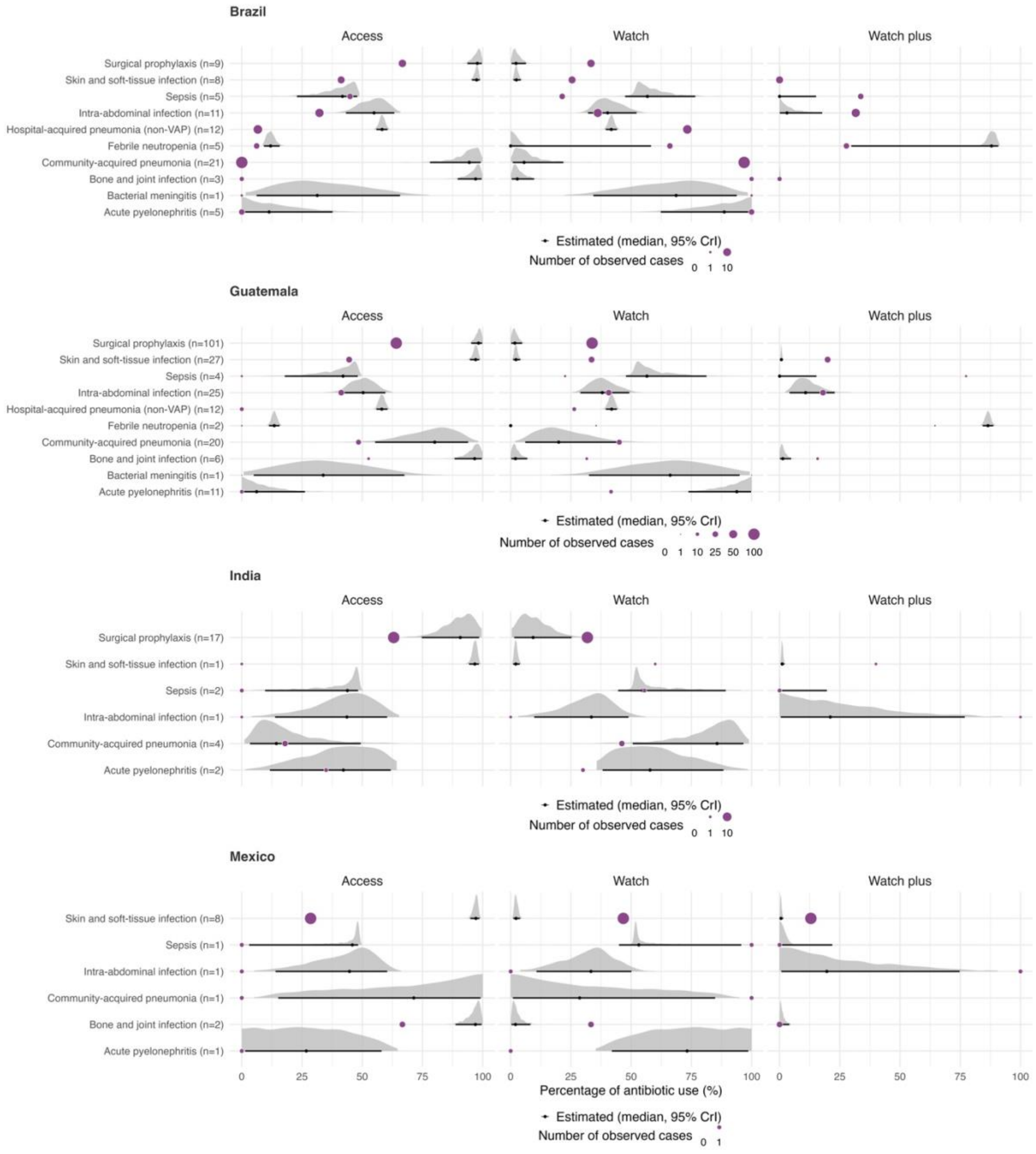

S Figure 16. Proportion of observed versus expected empiric antibiotic use out of the total use within each indication under Scenario 1 in patients > 12 years in 2021 in Brazil, Guatemala, India and Mexico. Purple coloured circular dots indicate the proportion of observed use within each indication. The ridge line plot represents the distribution of the proportion of the expected use. The total number of patients on empirical treatment used to estimate expected antibiotic use is shown in brackets (n).

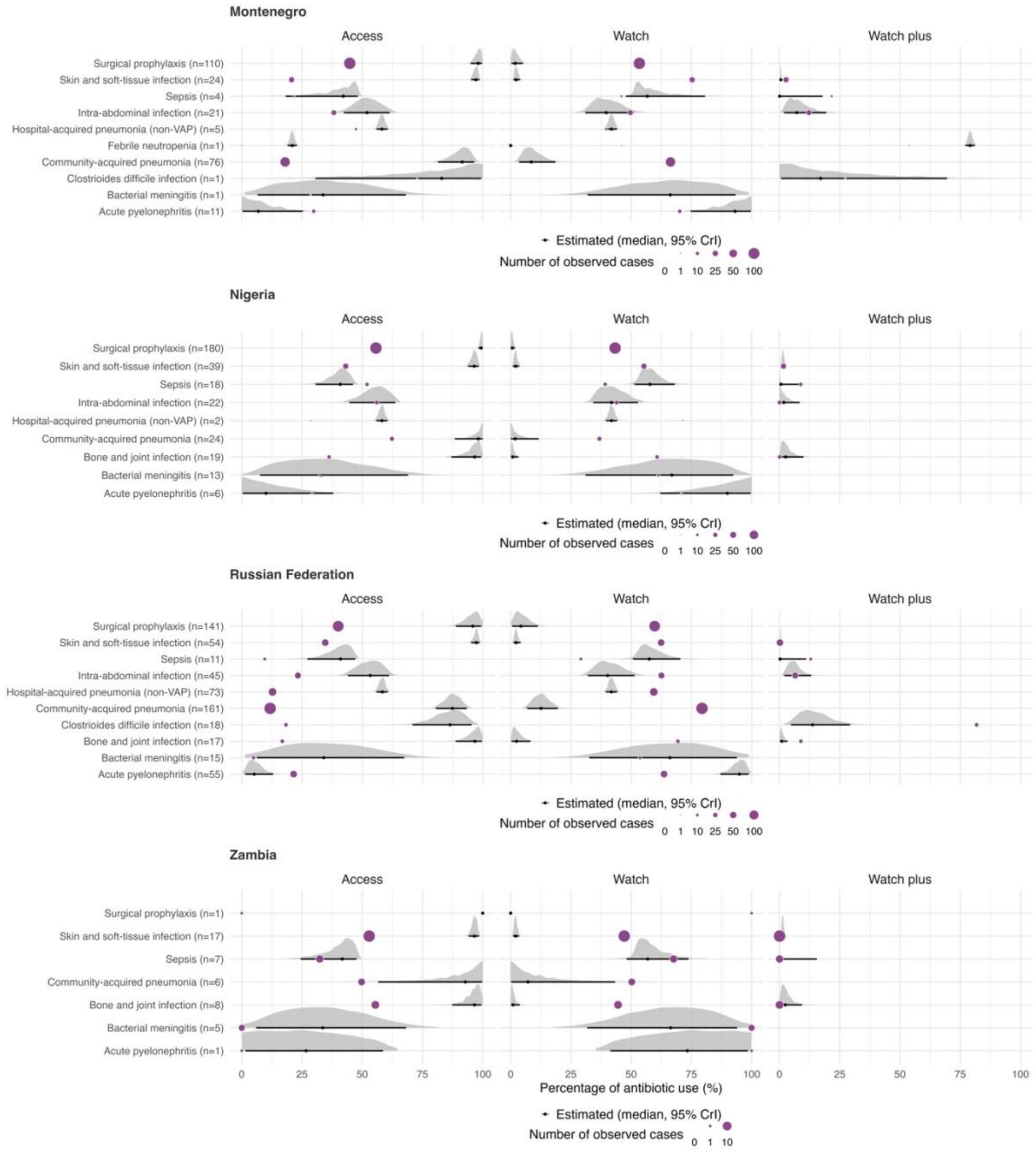

S Figure 17. Proportion of observed versus expected empiric antibiotic use out of the total use within each indication under Scenario 1 in patients > 12 years in 2021in Montenegro, Nigeria, Russian Federation and Zambia. Purple coloured circular dots indicate the proportion of observed use within each indication. The ridge line plot represents the distribution of the proportion of the expected use. The total number of patients on empirical treatment used to estimate expected antibiotic use is shown in brackets (n).

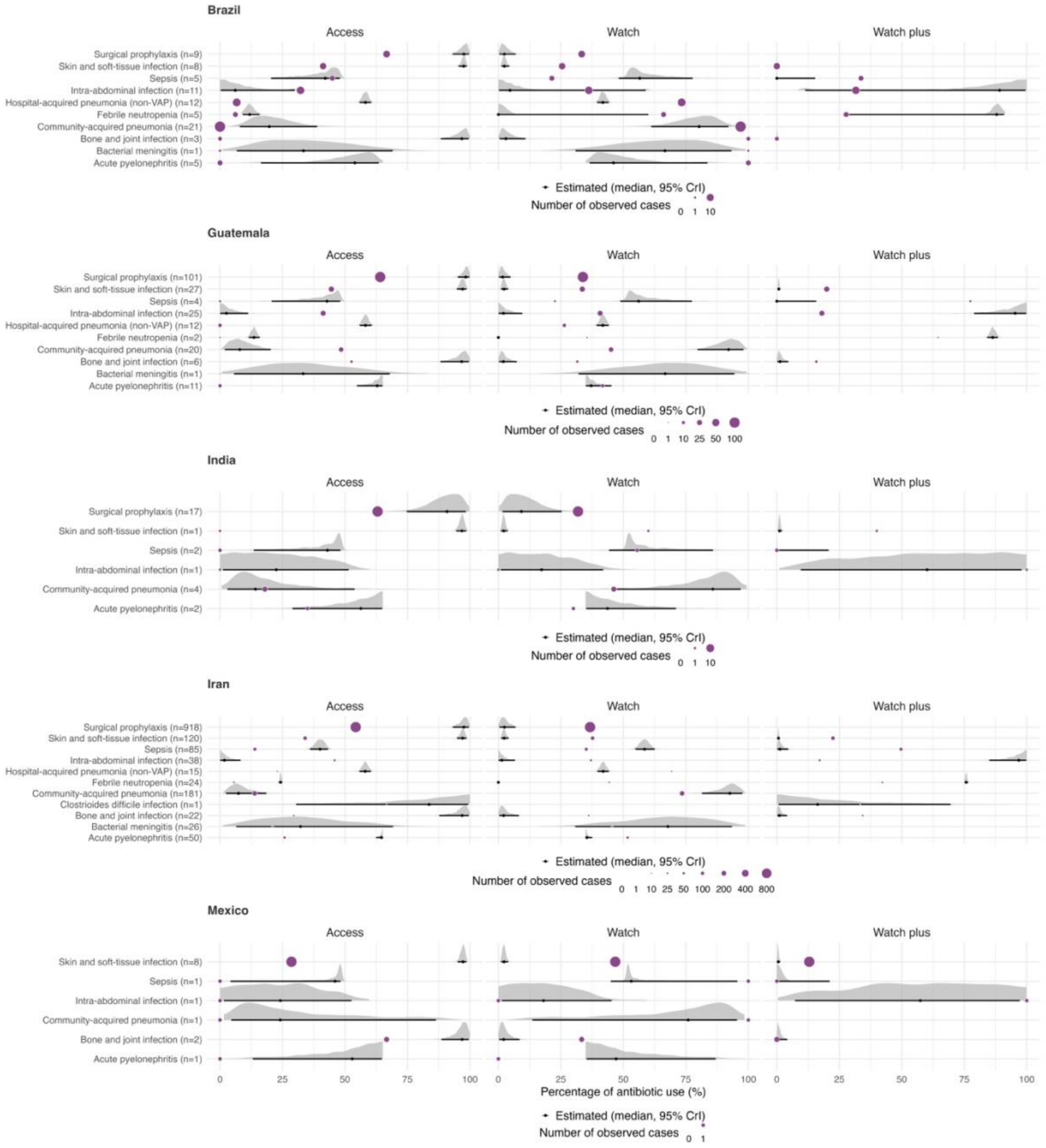

S Figure 18. Proportion of observed versus expected empiric antibiotic use out of the total use within each indication under Scenario 2 in patients > 12 years in 2021 in Brazil, Guatemala, India, Iran and Mexico. Purple coloured circular dots indicate the proportion of observed use within each indication. The ridge line plot represents the distribution of the proportion of the expected use. The total number of patients on empirical treatment used to estimate expected antibiotic use is shown in brackets (n).

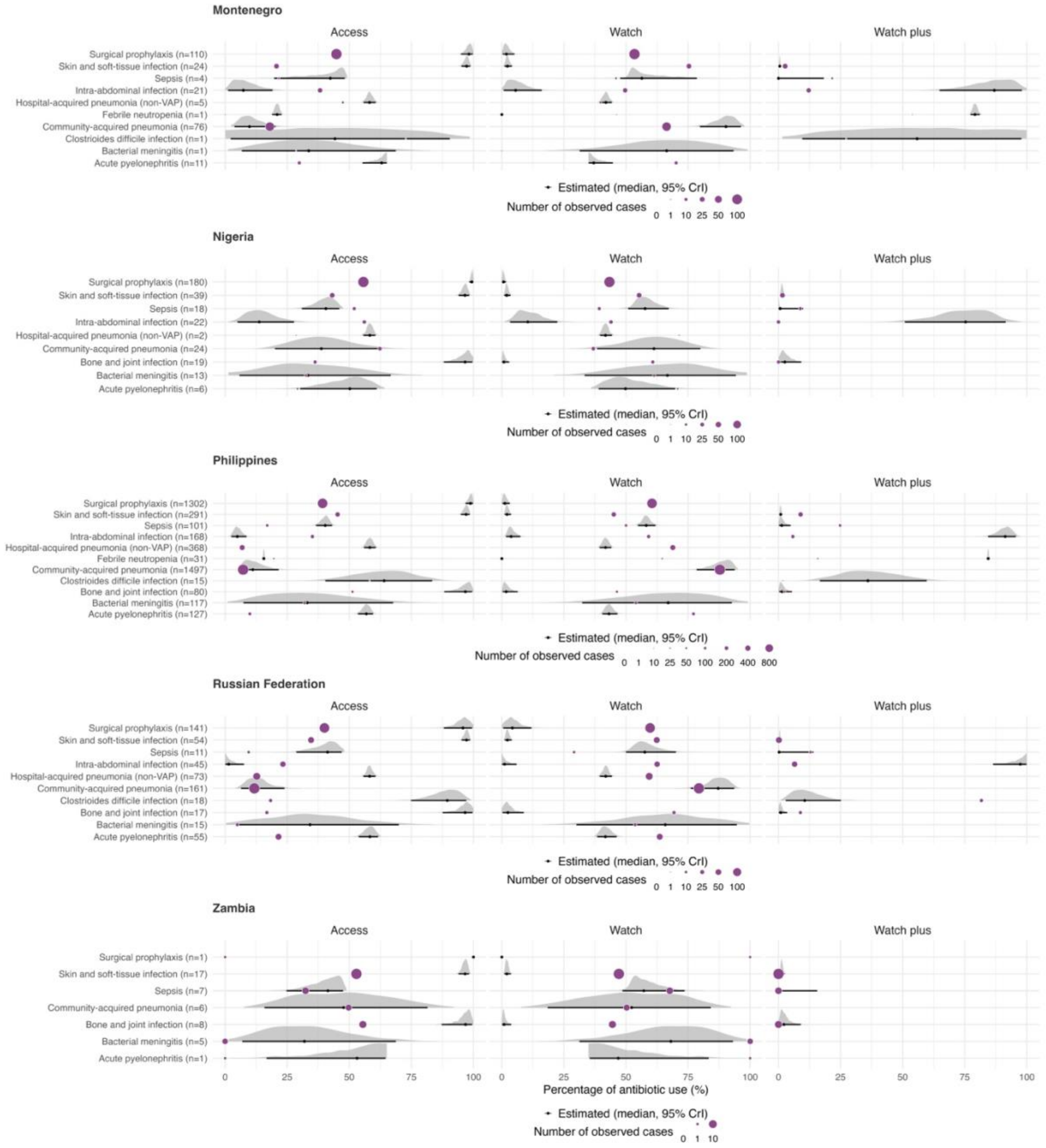

S. Figure 19. Proportion of observed versus expected empiric antibiotic use out of the total use within each indication under Scenario 2 in patients > 12 years in 2021in Montenegro, Nigeria, Philippines, Russian Federation and Zambia. Purple coloured circular dots indicate the proportion of observed use within each indication. The ridge line plot represents the distribution of the proportion of the expected use. The total number of patients on empirical treatment used to estimate expected antibiotic use is shown in brackets (n).

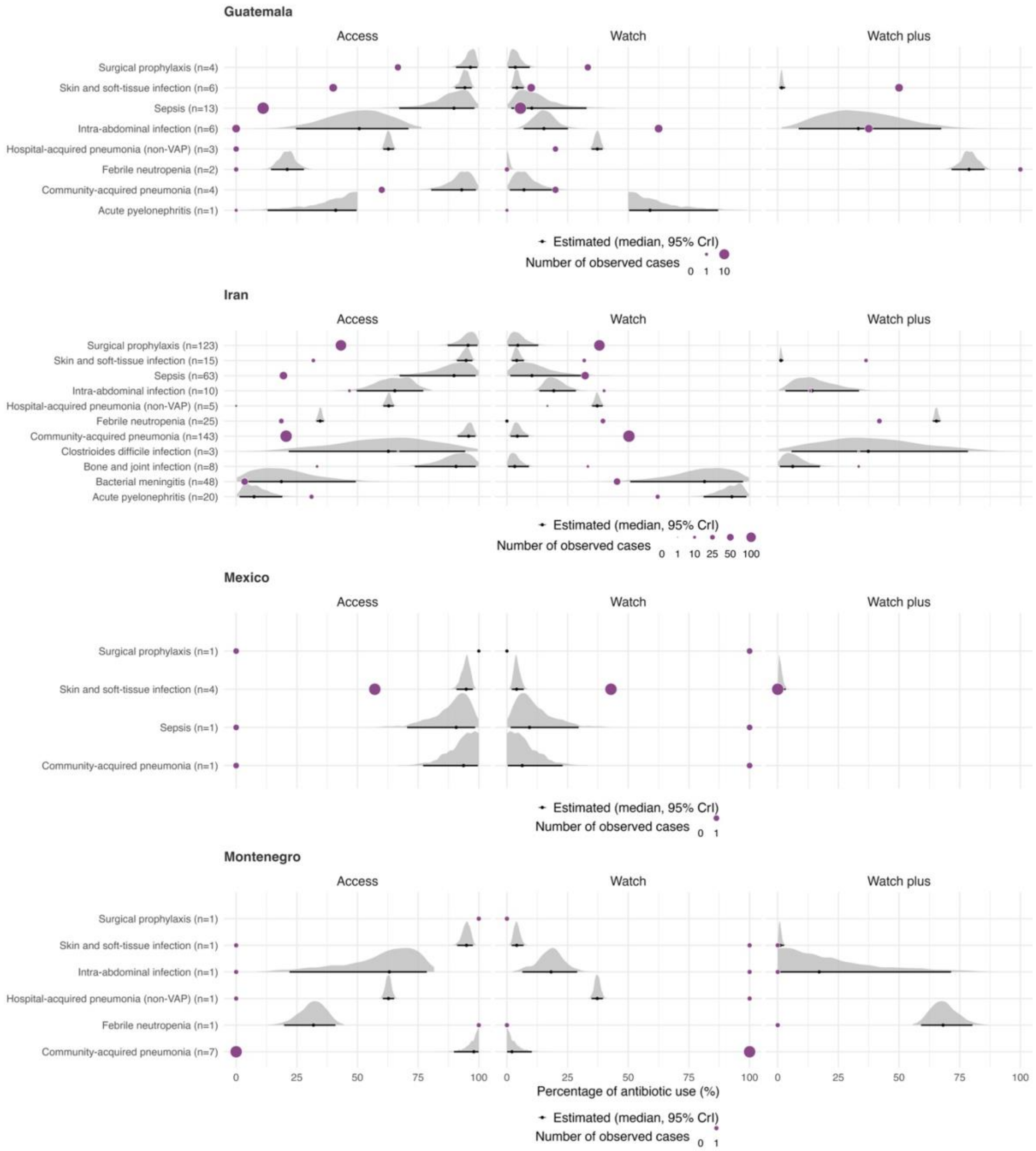

S Figure 20. Proportion of observed versus expected empiric antibiotic use out of the total use within each indication under Scenario 1 in patients ≤ 12 years in 2021in Guatemala, Iran, Mexico and Montenegro. Purple coloured circular dots indicate the proportion of observed use within each indication. The ridge line plot represents the distribution of the proportion of the expected use. The total number of patients on empirical treatment used to estimate expected antibiotic use is shown in brackets (n).

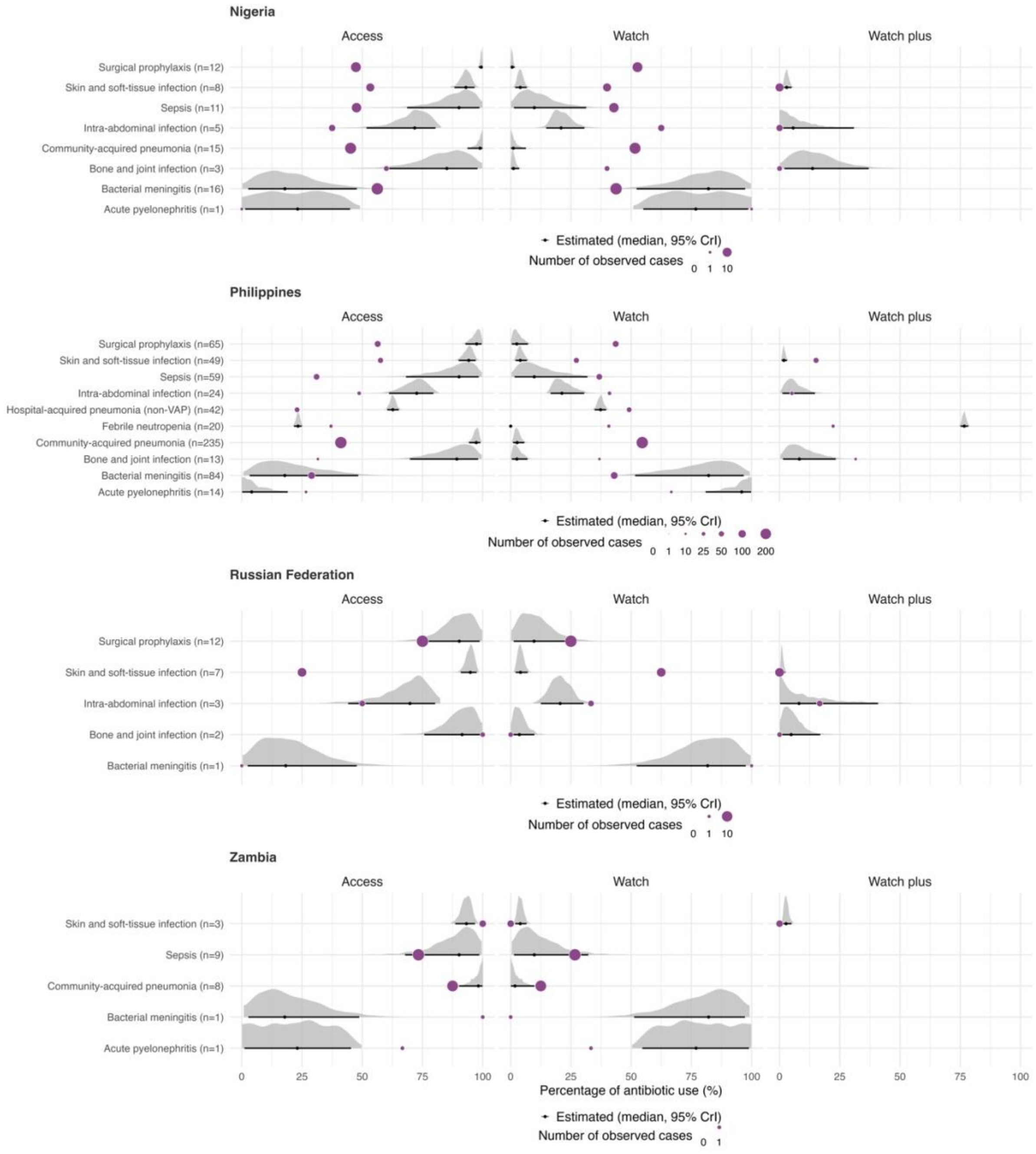

S Figure 21. Proportion of observed versus expected empiric antibiotic use out of the total use within each indication under Scenario 1 in patients ≤ 12 years in 2021in Nigeria, Philippines, Russian Federation and Zambia. Purple coloured circular dots indicate the proportion of observed use within each indication. The ridge line plot represents the distribution of the proportion of the expected use. The total number of patients on empirical treatment used to estimate expected antibiotic use is shown in brackets (n).

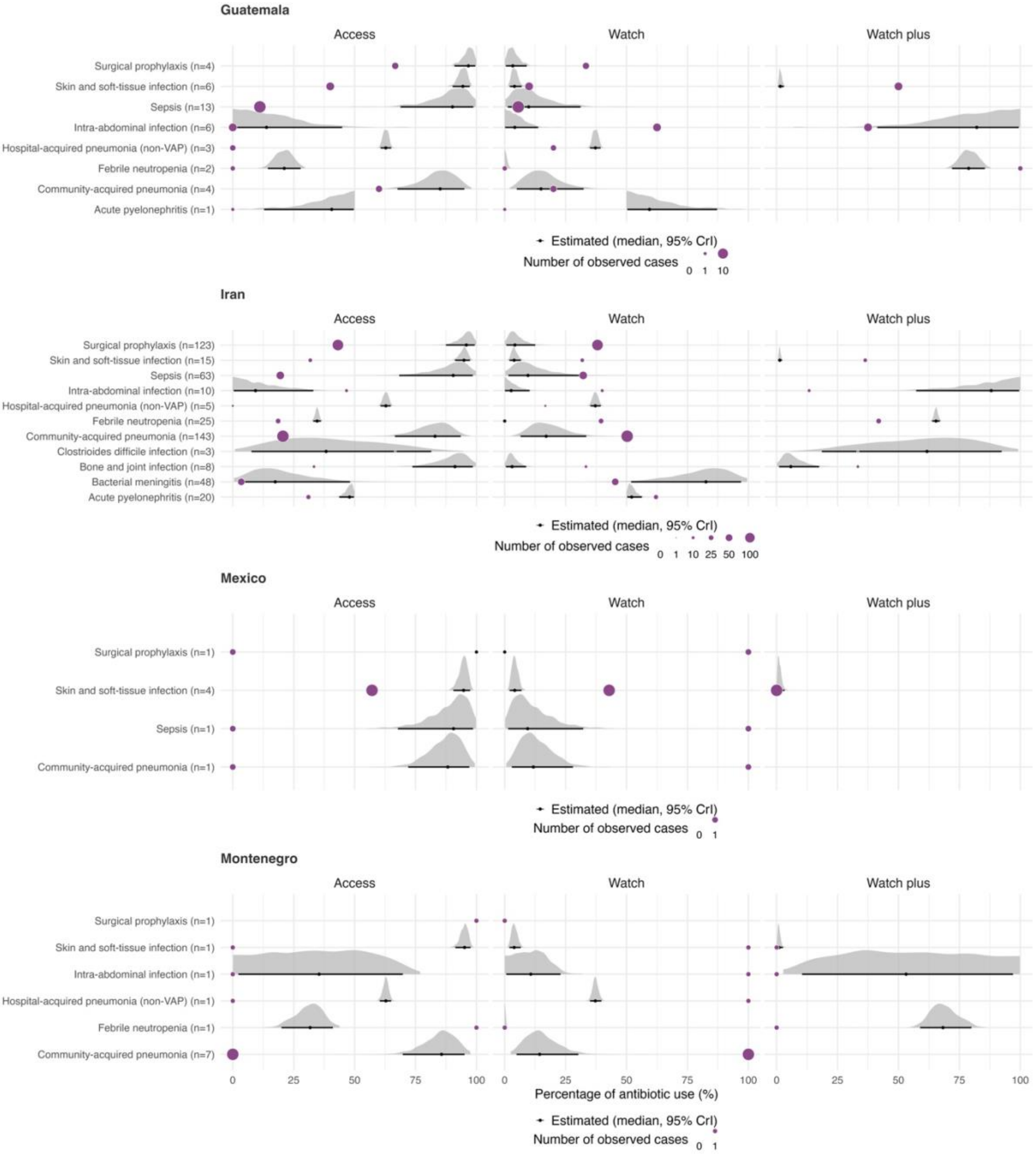

S Figure 22. Proportion of observed versus expected empiric antibiotic use out of the total use within each indication under Scenario 2 in patients ≤ 12 years in 2021in Guatemala, Iran, Mexico and Montenegro. Purple coloured circular dots indicate the proportion of observed use within each indication. The ridge line plot represents the distribution of the proportion of the expected use. The total number of patients on empirical treatment used to estimate expected antibiotic use is shown in brackets (n).

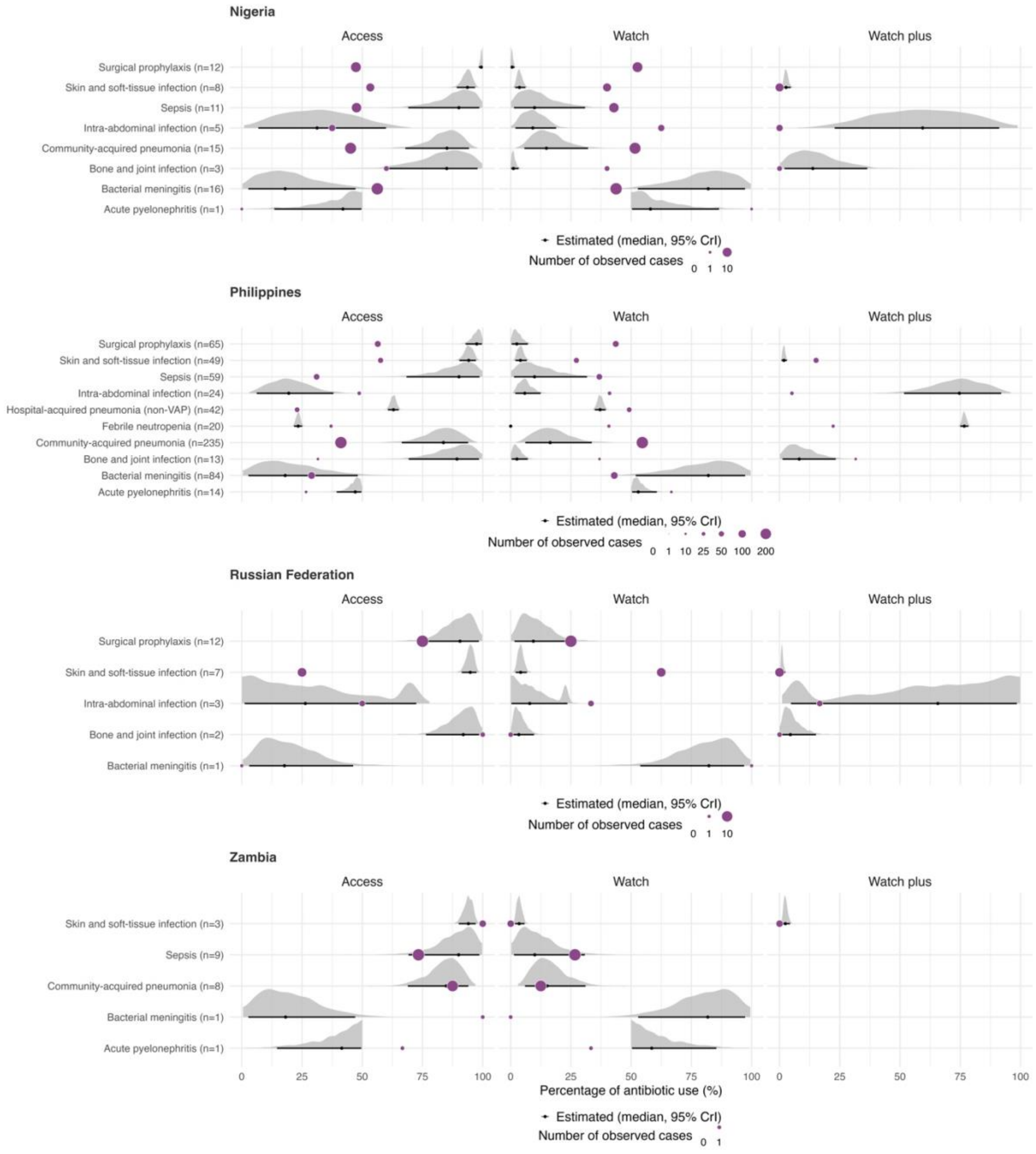

S Figure 23. Proportion of observed versus expected empiric antibiotic use out of the total use within each indication under Scenario 2 in patients ≤ 12 years in 2021in Nigeria, Philippines, Russian Federation and Zambia. Purple coloured circular dots indicate the proportion of observed use within each indication. The ridge line plot represents the distribution of the proportion of the expected use. The total number of patients on empirical treatment used to estimate expected antibiotic use is shown in brackets (n).

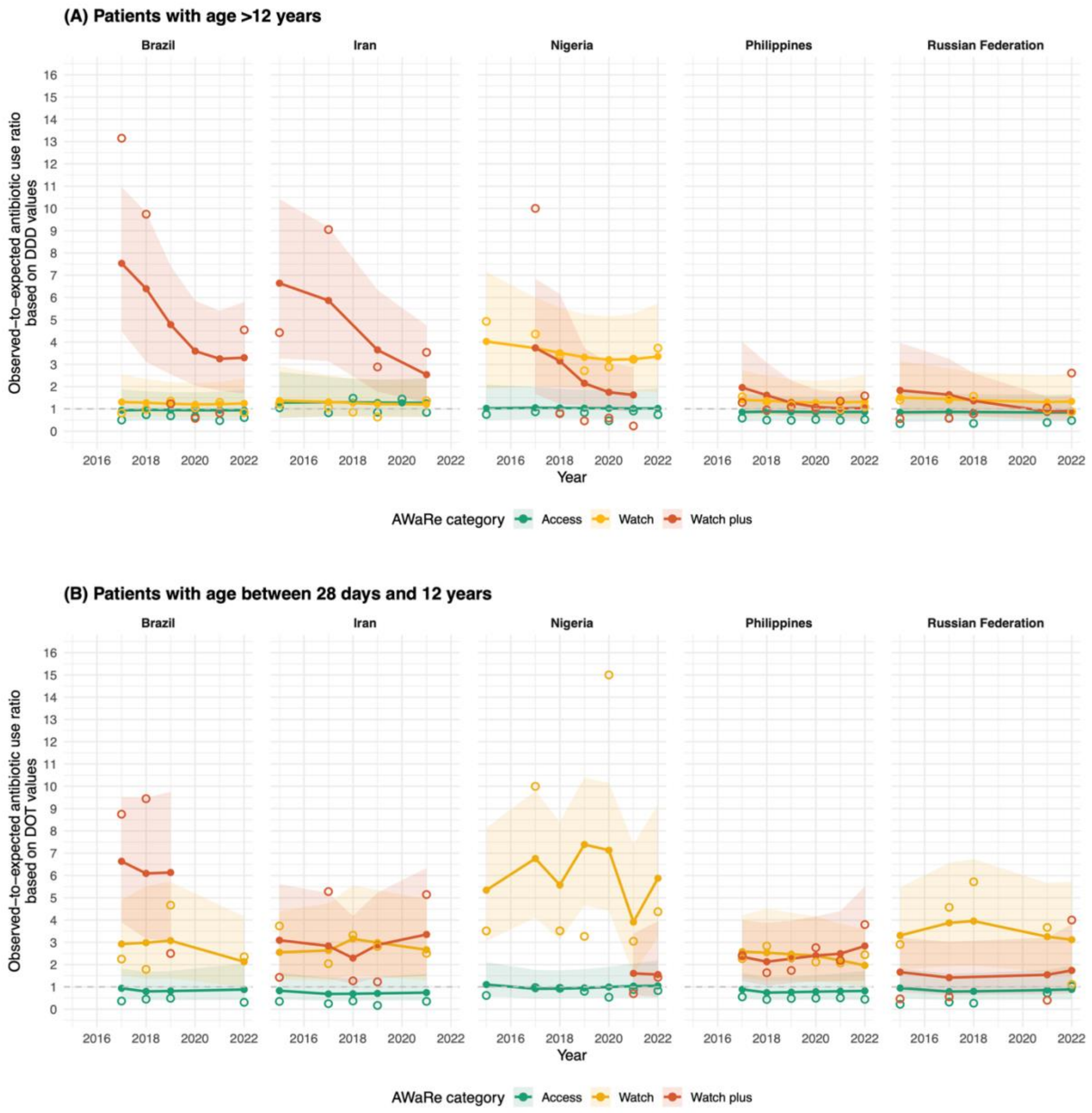

S Figure 24. Temporal changes in the observed-to-expected use ratio for the three AWaRe antibiotic groups among patients (A) aged >12 years and (B) 28 days to ≤12 years, based on data from five countries under Scenario 2 between 2015 and 2022. Hollow circles represent the calculated observed-to-expected use ratios, while solid circles represent estimates from a Bayesian multilevel model; shaded areas indicate the 95% credible intervals of the estimates. The horizontal dashed lines mark the value of 1, indicating that observed and expected antibiotic use (measured in DDD and DOT for patients aged >12 years and 28 days to ≤12 years, respectively) are equal.
